# Open-source computational pipeline automatically flags instances of acute respiratory distress syndrome from electronic health records

**DOI:** 10.1101/2024.05.21.24307715

**Authors:** Félix L. Morales, Feihong Xu, Hyojun Ada Lee, Helio Tejedor Navarro, Meagan A. Bechel, Eryn L. Cameron, Jesse Kelso, Curtis H. Weiss, Luís A. Nunes Amaral

**Affiliations:** Department of Engineering Science and Applied Mathematics, Northwestern University, Evanston, IL; Interdepartmental Biological Sciences Program, Northwestern University, Evanston, IL; Northwestern Institute on Complex Systems, Northwestern University, Evanston, IL; Medical Scientist Training Program, Northwestern University Feinberg School of Medicine, Chicago, IL; Department of Radiology, Emory University, Atlanta, GA; Department of Medicine, Endeavor Health, Evanston, IL; Division of Pulmonary and Critical Care Medicine, Endeavor Health, Evanston, IL; Department of Physics and Astronomy, Northwestern University, Evanston, IL; Department of Medicine, Northwestern University Feinberg School of Medicine, Chicago, IL; NSF-Simons National Institute for Theoretical and Mathematical Biology, Chicago, IL

**Author notes:** Corresponding Author: Luís A. Nunes Amaral.

## Abstract

Physicians, particularly intensivists, face information overload and decision fatigue, underscoring the need for automated diagnostic tools. Acute Respiratory Distress Syndrome (ARDS) affects over 10% of critical care patients, with over 40% mortality rate, yet is only recognized in 30-70% of cases in clinical settings. We present a reproducible computational pipeline that automates ARDS adjudication in retrospective datasets of mechanically ventilated adults, implementing the Berlin Definition via natural language processing and classification algorithms. We used labeled chest imaging reports from two hospitals to train an XGBoost model to detect bilateral infiltrates, and a labeled subset of attending physician notes from one hospital to train another XGBoost model to detect a pneumonia diagnosis. Both models achieve high discriminative performance on test sets—an area under the receiver operating characteristic curve (AUROC) of 0.88 for adjudicating bilateral infiltrates on chest imaging reports, and an AUROC of 0.87 for detecting pneumonia on attending physician notes. We integrated these models with rule-based components and validated the entire pipeline on a subset of healthcare encounters from a third hospital (MIMIC-III). We find a sensitivity of 93.5% in adjudicating ARDS — far surpassing the 22.6% ARDS documentation rate we found for this cohort — along with a false positive rate of 17.4%. We conclude that our reproducible, automated pipeline holds promise for improving ARDS recognition and could aid clinical practice through real-time EHR integration.

## Introduction

Physicians, especially intensivists, process large amounts of dispersed information from many patients^1^. This potential information overload poses serious risks to patient safety. Several studies have estimated that hundreds of thousands of fatalities per year may be due to medical errors^2–4^. While information overload is a challenge for humans, vast amounts of information become advantageous if used as an input for machine learning (ML) approaches. Recent advances in artificial intelligence, ML, and data science are enabling the development of protocols to extract knowledge from large datasets. However, some of those approaches lack interpretability and have been shown to be fragile (e.g., recent re-analysis of attempts to diagnose COVID-19 from chest X-ray images^5^).

In this study, we report on the development and validation of a computational pipeline to help physicians adjudicate acute respiratory distress syndrome (ARDS). ARDS, a syndrome of severe acute hypoxemia resulting from inflammatory lung injury^6^, is an ideal case for the development of a diagnostic aid tool. ARDS recognition requires physicians to synthesize information from multiple distinct data streams and determine whether it fits a standard definition. The clinically based Berlin Definition of ARDS criteria include quantitative data (PaO_2_/FiO_2_ ≤ 300 mm Hg), unstructured data (bilateral opacities on chest imaging), and subjective data (assessing for the presence of ARDS risk factors and heart failure)^6^. Despite ARDS’ high prevalence, morbidity, and mortality, prior research has shown that many patients with ARDS are not recognized by their treating physicians^7,8^. The poor recognition rate of ARDS^7^ is at least partially due to the difficulty in evaluating and integrating all the Berlin Definition criteria, which requires the physician to access laboratory data, chest images or radiology reports, other physicians’ notes, and echocardiographic data or reports, before they can apply the criteria to determine whether ARDS is present.

Under-recognition of ARDS plays an important role in under-utilization of evidence-based ARDS treatment (e.g., low tidal volume ventilation and prone positioning), even when physicians believe these interventions are warranted^9^. An automated approach to the adjudication of the ARDS diagnostic criteria has the potential to be a powerful aid to physician decision-making, leading to improved ARDS recognition and therefore improved ARDS management.

Previous studies have demonstrated some success in automating the recognition of individual ARDS diagnostic components using electronic health record (EHR) screening “sniffers”^10–12^. In addition, an ML algorithm to risk-stratify patients for ARDS using structured clinical data derived from the EHR was shown to have good discriminative performance^13^. Regarding automating the entire ARDS diagnostic algorithm, two studies have recently reported the implementation of keyword search (i.e. rule-based approach) in the EHR with validation conducted for 100 intensive care unit (ICU) admissions from a single period and from a single institution^14,15^. A third study recently reported on a computable Berlin Definition which employed a previously developed neural network approach to adjudicate chest imaging reports restricted to patients with a single, known ARDS risk factor (COVID-19), with promising performance (93% sensitivity, 92% specificity)^16^. However, no study has so far succeeded in simultaneously automating the entire sequence of steps required by the Berlin Definition of ARDS reproducibly, and testing the discriminative performance of the tool on a multi-hospital population of critically ill patients who received invasive mechanical ventilation. Our study addresses this gap in a manner that overcomes physician under-recognition of ARDS.

## Data and Methods

### Cohort Data Collection

We obtained a waiver of informed consent; the study was approved by the Institutional Review Boards of Northwestern University (STU00208049) and Endeavor Health (EH17-325). We collected data from three patient cohorts for model training and evaluation: Hospital A (2013), Hospital A (2016), and Hospital B (2017-18), where A and B refer to two distinct and unaffiliated hospitals and the number in parentheses refers to the period over which data was collected. In addition, we obtained data from the Medical Information Mart for Intensive Care (MIMIC-III) database^17–19^. MIMIC-III is a large, single-center database of critically ill patients at a tertiary care medical center. It includes all the components necessary to identify ARDS—and therefore apply our pipeline—, and it is freely available. For naming consistency, we denote the subset of MIMIC-III patients we consider as MIMIC (2001-12).

For all four cohorts, patient admissions (i.e., healthcare encounters, hereafter denoted encounters) were included if they were at least 18 years old; were admitted to an adult ICU; and had acute hypoxemic respiratory failure requiring intubation and invasive mechanical ventilation (at least one recorded PaO_2_/FiO_2_ ≤300 mm Hg while receiving positive-end expiratory pressure ≥ 5cm H_2_O)^8^. Table 1 summarizes the data available for the four cohorts.

**Table 1.** Availability of data and labels by hospital. Hospital A, Hospital B, and MIMIC refer to three distinct and unaffiliated hospitals and the number in parentheses refers to the period over which data was collected.

| Source | Report type | Number | % bilateral infiltrates | Rater IDs |
| --- | --- | --- | --- | --- |
| Hospital A (2013) | Chest imaging (CXR and CT) reports | 5,958 reports (800 encounters) | 57% | 1 |
| Hospital A (2013) | Attending physician notes | 12,582 notes (790 encounters) |  |  |
| Hospital A (2013) | Adjudicated Attending physician notes | 2,034 notes (400 encounters) |  | 1, 5 |
| Hospital A (2013) | Echocardiogram reports | 1,006 reports (681 encounters) |  |  |
| Hospital A (2016) | Chest imaging (CXR and CT) reports | 6,040 reports (749 encounters) | 44% | 1 |
| Hospital B (2017-18) | Chest imaging (CXR and CT) reports | 625 reports (90 encounters) | 34% | 1, 2, 3, 4 |
| MIMIC (2001-12) | Chest Imaging (CXR and CT) reports | 975 reports (100 encounters) | 22% | 1,6 |
| MIMIC (2001-12) | Attending physician notes | 887 notes (100 encounters) |  | 1,6 |
| MIMIC (2001-12) | Echocardiogram reports | 89 reports (100 encounters) |  | 1,6 |

### Hospital A (2013)

We previously characterized a cohort of 943 encounters, which we denote here as Hospital A (2013), who met the above inclusion criteria at a single academic medical center between June and December 2013^8^. We collected the following data: all PaO_2_/FiO_2_ ratios (hereafter, PF ratios); the unstructured text of all radiologist reports for chest imaging (radiographs and CT scans), critical care attending physician notes, and echocardiogram reports; and B-type natriuretic peptide (BNP) values obtained from hospital admission to the earliest of extubation, death, or discharge. Data were reviewed by study personnel to determine whether each individual Berlin Definition criterion was present, and whether all criteria taken together were consistent with a diagnosis of ARDS^8^.

We collected 5,958 chest imaging reports from 800 Hospital A (2013) encounters. Study personnel adjudicated whether these chest imaging reports described bilateral infiltrates consistent with the Berlin Definition. In case of disagreement, a senior critical care physician served as tiebreaker. At the end of this process, 57% of these chest imaging reports were labeled as describing bilateral infiltrates consistent with the Berlin Definition^6^. We only had access to this tie-breaking label. We developed our machine learning (ML) approach to bilateral infiltrate adjudication using these Hospital A (2013) reports.

For 790 of the 800 Hospital A (2013) encounters with a chest imaging report, we also had at least one attending physician note. We collected 12,582 attending physician notes for these encounters, of which 2,034 notes from a subset of 400 encounters were labeled by study personnel for the presence of ARDS risk factors (e.g., pneumonia, sepsis, aspiration, etc.)^6^. We used this labeled subset of 2,034 notes to develop our ML and regular expression (regex) approach for finding ARDS risk factors and heart failure language in attending physician notes.

We collected 1,006 echocardiogram (echo) reports from 681 Hospital A (2013) encounters. Study personnel from a prior analysis^8^ text-matched and adjudicated each echo report for the presence or absence of left ventricular ejection fraction < 40%, cardiopulmonary bypass at time of echo, left ventricular hypertrophy, left atrial dimension > 4cm or left atrial volume index > 28 mL/m^2^, and Grade II or III diastolic dysfunction. We also included 35 BNP values for 32 encounters in Hospital A (2013). We used echo reports and BNP values to develop our objective heart failure rule-out approach^8^.

Since identifying Berlin Definition-consistent bilateral infiltrates is the most challenging task^12,20^ in our computational pipeline, we analyzed the chest imaging reports from two additional cohorts to test our computational pipeline, Hospital A (2016) and Hospital B (2017-18).

### Hospital A (2016)

The Hospital A (2016) cohort comprises 749 encounters admitted during 2016 at the same medical center as the Hospital A (2013) cohort and meeting the same inclusion criteria. We collected 6,040 chest imaging reports (radiographs and CT scans) for these encounters. Like Hospital A (2013), we only had access to the tie-breaking labels for these reports, adjudicated 44% of Hospital A (2016) reports as describing bilateral infiltrates. We developed our machine learning (ML) approach to bilateral infiltrate adjudication using these Hospital A (2016) reports.

### Hospital B (2017-18)

The Hospital B (2017-18) cohort comprises 90 encounters admitted to a different, unaffiliated, medical center in 2017–2018 and meeting the same inclusion criteria as Hospital A (2013) and Hospital A (2016). We collected 625 chest imaging (radiographs and CT scans) reports for these 90 encounters. The tie-breaking critical care physician adjudicated 34% of these chest imaging reports as describing bilateral infiltrates. Depending on the experiments (see below), we used these reports from Hospital B (2017-18) to develop our ML models for bilateral infiltrate adjudication, or to evaluate the robustness of XGBoost to changes in the training data.

### MIMIC (2001-12)

We identified the set of encounters in the MIMIC-III dataset who satisfied the inclusion criteria used to develop Hospital A (2013). This resulted in a set comprising 3,712 encounters. We then used our pipeline to adjudicate the presence or absence of ARDS for all those encounters, and randomly selected a balanced cohort comprising 100 encounters, which we denote as the MIMIC (2001-12) cohort. Each of the encounters in MIMIC (2001-12) was adjudicated by one experienced critical care physician and one internal medicine physician for whether each Berlin Definition criterion was present, and whether all criteria taken together were consistent with a diagnosis of ARDS. We note that both reviewers were blinded to the individual encounter adjudications by the pipeline, although they did know that the ARDS split among the 100 encounters was balanced. This cohort, with blind physician adjudications, is publicly available at Northwestern’s ARCH database.

The records of MIMIC (2001-12) encounters included 975 chest imaging (radiographs and CT scans) reports, 887 attending physician notes, and 89 echocardiogram (echo) reports. The critical care physician adjudicated 22.3% of these chest imaging reports as describing bilateral infiltrates consistent with the Berlin Definition^6^. The same individual also labeled 887 attending physician notes for the presence of ARDS risk factors^6^, heart failure language, and whether ARDS was mentioned in the note (alongside mentions of Acute Lung Injury given that this dataset predates the publication of the Berlin Definition). The critical care physician also labeled 89 echo reports for the presence or absence of left ventricular ejection fraction < 40%, cardiopulmonary bypass at time of echo, left ventricular hypertrophy, left atrial dimension > 4cm or left atrial volume index > 28 mL/m^2^, and Grade II or III diastolic dysfunction^8^. We used these adjudicated datasets to evaluate: (1) the performance of our implemented ML models, and (2) the performance of the entire pipeline on a publicly available dataset.

## Methods

### Adjudication of bilateral infiltrates from chest imaging reports

#### Overview

We used labeled chest imaging reports from Hospital A (2013), Hospital A (2016), and Hospital B (2017-2018) to develop a machine learning (ML) approach to adjudicating bilateral infiltrates in chest imaging reports. We will refer to this data as the “development set” — it comprises a total of 12,623 chest imaging reports of which 6272 (49.7%) reports were labeled positive for bilateral infiltrates.

In addition, we tested the robustness of the XGBoost classifier to changes in the training data by training two separate models: one trained on chest imaging reports from Hospital System A (2013), and another on chest imaging reports from Hospital System A (2016). We did not carry these models forward for use in the ARDS adjudication pipeline. Finally, we used all the 975 chest imaging reports from MIMIC (2001-12) as our test set for the Bilateral Infiltrates Model.

#### Feature engineering

We preprocessed chest imaging reports by removing patient and clinician identification information, irrelevant sections (e.g. technique, indication, history, etc.), and ‘non-informative’ words. In selecting informative vs. non-informative words, we used the same inclusion and exclusion language criteria we previously developed to address limitations in Berlin Definition specificity^8^. We then tokenized the remaining sections — i.e., separated the text into sets of unigrams and bigrams — and prepared the data for use of a “bag of words” approach — i.e., vectorized these tokens according to their counts in the imaging reports. When training an ML model on a given corpus, we used the 200 most frequently appearing tokens across the imaging reports from a given corpus as model features.

#### Training of models for the adjudication of bilateral infiltrates

We compared the performance of four different classifiers — decision trees, logistic regression, random forest, and extreme gradient boosting ‘XGBoost’^21^ — on the development set (see *Overview*). We implemented a <u>nested</u> cross-validation strategy where the <u>outer loop</u> consisted of a 5-fold cross-validation and, within each <u>inner loop</u>, each fold’s training set was used to tune that model’s hyperparameters using a 3-fold cross-validation strategy. We chose a 3-fold cross validation strategy for the inner loop, instead of the 5-fold used for the outer loop, due to the computational cost of running nested cross-validation on 12 thousand records.

#### Preventing data leakage

Unless otherwise noted, all cross-validation strategies used healthcare encounters, not individual reports. We split the reports this way to avoid “data leakage” — that is, having chest imaging reports from the same encounter appear in both the training and validation data. Thus, we ensured all reports from a given encounter can only be found on either the training or validation data (but not both). We also used nested cross-validation to prevent data leakage, as this avoids tuning hyperparameters on validation data.

#### Hyperparameter tuning

We used the Bayesian optimization implemented in *hyperopt* package (v.0.2.7)^22^ for Python (v.3.10.12) to perform hyperparameter tuning. For each model trained, we performed cross-validation to obtain the mean log loss for each hyperparameter combination considered. We then selected the optimal combination of hyperparameters as the one yielding the lowest cross-validation mean log loss after at least 100 iterations. However, we employed an early-stopping rule to avoid running fruitless iterations.

#### Model comparison and feature importance

We used 95% confidence intervals to compare receiver operating characteristic (ROC) curves, areas under the ROC curves (AUROCs), and calibration curves across different models. For obtaining feature/token importance during training, we employed the default “importance” method that version 1.1.3 of the scikit-learn package implements for decision tree, logistic regression, and random forest, and version 1.7.4 of *xgboost* package for XGBoost. For decision trees and random forests, feature importance corresponds to the mean decrease in Gini impurity; for logistic regression, importances correspond to the mean value of coefficients in the fitted linear equation; and for XGBoost, the importance corresponds to the mean gain in predictive performance obtained by including a particular feature in the trees. We additionally used Shapley-additive explanations (SHAP; v.0.42.0) values to obtain the feature/token importance on test sets.

#### Model testing for adjudication of bilateral infiltrates

Model selection resulted in XGBoost being chosen as the best performing classifier for the task of adjudicating bilateral opacities in chest imaging reports. To assess the robustness of XGBoost to different training sets, we tuned hyperparameters and then trained XGBoost models on all chest imaging reports from Hospital A (2013) and Hospital A (2016), separately. We then tested each of the two models on the two other chest imaging corpora *the model had not yet seen* by comparing the mean AUROC values after 5-fold cross validation. We used 10 resamples with replacement (bootstrap) to obtain 95% confidence intervals for the estimates of the mean AUROC values. This test also allowed us to scope out the generalizability of our approach.

However, we finally tested XGBoost’s generalization by tuning its hyperparameters and then training it on the full development set. We then applied this model to the 975 chest imaging reports from MIMIC (2001-12), employing a similar cross-validation and bootstrapping approach as described above for testing XGBoost’s robustness. It is this model that we carried forward for use in the adjudication pipeline, and that we’ll refer to as the “Bilateral Infiltrates Model”.

We also evaluated the inter-rater disagreement rate for chest imaging reports from MIMIC (2001-12). For this purpose, we obtained two independent adjudications (one critical care physician and one internal medicine physician) for the 975 reports, and split imaging reports into three groups according to whether both physician raters agreed on a No, agreed on a Yes, or disagreed. For each group, we then calculated the mean output probability by the Bilateral Infiltrates Model. In addition, we also split imaging reports into three groups according to the Bilateral Infiltrates Model’s output probabilities. For each group, we then calculated the fraction of imaging reports for which each independent rater disagreed with the Bilateral Infiltrates Model’s adjudications.

### Adjudication of risk factors from physician notes

#### Overview

We used 2,034 adjudicated or labeled attending physician notes from Hospital A (2013) to develop our approaches to adjudicate ARDS risk factors and/or heart failure. However, not every one of the 2034 notes was labeled for every risk factor. For a full list of risk factors and heart failure language available for Hospital A (2013) notes, as well as how many notes were labeled for each, see SI: Regular expressions list 1 and Table S1. We did not have labeled attending physician notes available for Hospital System A (2016) or Hospital B (2017-2018). However, we had 887 labeled attending physician notes from MIMIC (2001-12) available, and we used them as our test set. Unlike notes from Hospital A (2013), every note from MIMIC (2001-12) was labeled by a critical care physician for the following criteria: pneumonia, aspiration, burns, pancreatitis, pulmonary contusion, sepsis, trauma, vasculitis, and cardiac surgery.

#### Capturing risk factors with regular expressions

The Berlin Definition of ARDS requires the presence of at least one risk factor — e.g., pneumonia, sepsis, shock, inhalation, pulmonary contusion, vasculitis, drowning, drug overdose — within seven days of non-cardiogenic acute respiratory failure. We preprocessed labeled attending physician notes from Hospital A (2013) to remove identifiable information from the text of these notes. We then used regular expressions (*regex* v2022.10.31) to match keywords related to risk factor and heart failure language that had labels available in Hospital A (2013) (see SI: Regular expression list 1, for a complete list of risk factors and regex patterns used to capture them). To validate this strategy, we ensured that this regex approach captured 100% of the notes that had a positive label for a particular risk factor (or close to 100% as possible). We also corrected common spelling errors on important keywords, such as ‘pneunonia’, ‘spetic’ or ‘cardigenic’.

#### Adjudication of risk factors with XGBoost - Feature engineering

Adjudicating risk factors requires more than simple keyword matching in physician notes. For instance, “patient is unlikely to have pneumonia” should not be classified as pneumonia. To address this, we applied an XGBoost approach for risk factors that had sufficient data (see below), mirroring our method for bilateral infiltrates unless specified otherwise. Lacking predefined inclusion/exclusion criteria for physician notes, we extracted a 200-character window around matched keywords (100 characters before and after) using regex. These text segments were then tokenized and vectorized into token frequency matrices, as done for the Bilateral Infiltrates Model.

#### Adjudication of risk factors with XGBoost - Risk factor/model selection

Using the vectorized tokens, we trained XGBoost models for a select group of risk/heart failure factors. We only used a select group since not all risk factors were amenable to ML approaches: we chose risk factors that had more than 100 notes adjudicated in Hospital A (2013) notes corpus and had relatively balanced yes/no proportions after regex-matching (between 33% and 66%, see SI Table 1). This resulted in the use of 1409 labeled attending notes from 337 encounters for XGBoost model development. It also resulted in only trying an XGBoost approach for the following risk factors or heart failure language: pneumonia, aspiration, congestive heart failure, and sepsis. In other words, we did not use all 2,034 labeled records from Hospital A (2013) to train each of the models since not every record had an annotation or a keyword for a given risk factor. For instance, only 636 notes in Hospital A (2013) included an annotation for pneumonia, whereas we were able to match 955 notes for pneumonia using regex. Therefore, our training dataset for each of the four models consisted of all notes that were regex-captured for that particular risk factor (e.g. 955 notes for the Pneumonia Model; see SI Table 1 for number of notes used to train each model). For notes that were regex-captured but did not have a label, we imputed the label as ‘No’, or zero (see SI Table 1 for label proportions after this imputation step).

We employed a similar nested cross-validation strategy to the one pursued for the adjudication of chest imaging reports. Notably, we also split the adjudicated notes into train and test sets by encounter, not note, to prevent data leakage. However, since we only had up to 1,409 notes for training, this task was not as computationally expensive as it was for adjudicating chest imaging reports. Therefore, we used 100 resamples instead of 10 and always employed 5-fold cross validation for hyperparameter tuning.

#### Adjudication of risk factors with XGBoost - model testing

Model selection yielded pneumonia as the risk factor best suited to be adjudicated by an XGBoost classifier. Therefore, we tested this “Pneumonia Model” by tuning XGBoost’s hyperparameters and then training it on the 955 attending physician notes that were captured by the pneumonia regex pattern. We then tested this Pneumonia Model on the 790 attending physician notes from MIMIC (2001-12) that were captured by the same pneumonia regex pattern, employing a similar cross-validation and bootstrapping approach as described above for testing the Bilateral Infiltrates Model. It is this model which we carried forward for use in the pipeline.

#### Adjudication of risk factors with regular expressions

We used an XGBoost model only for adjudicating pneumonia, applying a regex approach for other risk factors and heart failure criteria. We implemented two criteria to guide the use of regex: first, if a risk factor or heart failure criterion was labeled ‘yes’ in over 80% of notes, we applied the same regex pattern. Second, for more balanced labels, we refined the regex by adding “exclusion” words or phrases to reduce false positives (Regular Expression List 2). This set of exclusions was developed from notes at Hospital A (2013) and targeted risk factors suitable for refinement.

Finally, we tested both regex approaches on MIMIC (2001-12) notes labeled for the same risk factors and heart failure criteria available in Hospital A (2013): aspiration, burns, pancreatitis, pulmonary contusion, sepsis, trauma, and vasculitis. Only regex patterns meeting the predefined criteria for any hospital cohort were incorporated into the pipeline.

### Design of ARDS adjudication pipeline: chaining Berlin Definition steps

Each of the steps outlined above automates the adjudication of specific criteria in the Berlin Definition, with the modifications specified previously^8^. We integrate the different criteria to build a single, automated ARDS adjudication pipeline. The ARDS adjudication pipeline first flags encounters with at least one hypoxemia measurement (i.e., one instance of PF ratio ≤ 300 mm Hg while PEEP ≥ 5 cm H20). These PF ratios were precalculated for Hospital A (2013) and MIMIC (2001-12), see the Supplementary Information for more details.

After flagging hypoxemic entries, the pipeline uses the predictions of the Bilateral Infiltrates Model to adjudicate presence of bilateral infiltrate language in chest imaging reports. Upon settling these two criteria, the pipeline flags whether the hypoxemia record and the report consistent with bilateral infiltrates have timestamps within 48 hours of each other (which we term “qualified hypoxemia”). In addition, at this step the pipeline ensures that the hypoxemia record was taken at or after intubation. We chose a 48-hour window to be consistent with previous choices for manually adjudicating ARDS on Hospital A (2013)^8^. Since the Berlin Definition does not prescribe a time window, prior work settled on 48 hours to maximize recognition of ARDS retrospectively. Nevertheless, we observe that 75% of chest imaging reports from MIMIC (2001-12) had timestamps that were within 15.2 hours of a hypoxemia timestamp (Fig. S1).

Next, the pipeline uses the predictions of the Pneumonia Model to adjudicate pneumonia on all notes and uses regex to flag presence of other ARDS risk factors, heart failure language, and indicators of cardiogenic and noncardiogenic language. While we developed regex patterns for all risk factors and heart failure language available, we only included risk factors and heart failure language whose regex patterns fulfilled the criteria laid out in the previous section. As a result, for Hospital A (2013), we used a regex approach to only adjudicate *sepsis, shock and its “cardiogenic” qualifier, inhalation, pulmonary contusion, burns, vasculitis, drowning, and overdose*. In addition, for MIMIC (2001-12), we used a regex approach to also adjudicate *aspiration and pancreatitis.* Finally, the pipeline flags whether an attending physician note has a timestamp that falls between one day prior to and seven days after the latter timestamp of any of the qualifying hypoxemia-bilateral infiltrates pairs.

Once these annotations are integrated, the pipeline proceeds to adjudicate whether an ARDS adjudication is warranted. If any risk factor is identified in this time window for an encounter, regardless of whether heart failure is identified, the pipeline adjudicates the encounter as positive for ARDS. If no risk factors are identified, but only heart failure is identified, the pipeline adjudicates the encounter as negative for ARDS.

For all other encounters not meeting the risk factor or heart failure language criteria described above, the pipeline sends the encounter for objective heart failure assessment^8^. This assessment is done sequentially instead of by flagging: If any encounter had BNP greater than 100 pg/mL (an indicator of heart failure), the pipeline adjudicates those encounters as negative for ARDS. The pipeline then considers the remaining encounters for each subsequent criteria, adjudicating them as negative for ARDS if the encounter had any of the following: left ventricular ejection fraction < 40%, cardiopulmonary bypass found in echocardiogram report, or at least two of the following present in the echocardiogram report: (i) left atrial diameter > 4 cm or left atrial volume index > 28 mL/m2, (ii) left ventricular hypertrophy, or (iii) Grade II or III diastolic dysfunction.

Any encounter that is not adjudicated negative for ARDS after the objective heart failure assessment step is adjudicated as positive for ARDS. That is, the pipeline adds these encounters to those adjudicated as positive for ARDS via risk factor identification.

## Results

### Adjudication of bilateral infiltrates

Figure 1a shows the receiver operating characteristic (ROC) curves for the XGBoost model applied to chest imaging reports from the development set. We quantify model predictive performance using the areas under the ROC curves (AUROCs). We observe that once hyperparameters for each model are optimized, all models trained on chest imaging reports from the development set achieve AUROCs of at least 0.85 (Decision tree: AUROC = 0.87, 95%CI = [0.85, 0.89]; logistic regression: AUROC = 0.91, 95%CI = [0.91, 0.92]; random forest: AUROC = 0.93, 95%CI = [0.93, 0.93]; XGBoost: AUROC = 0.93, 95%CI = [0.93, 0.94]) (Fig. 1b).

**Figure 1.**
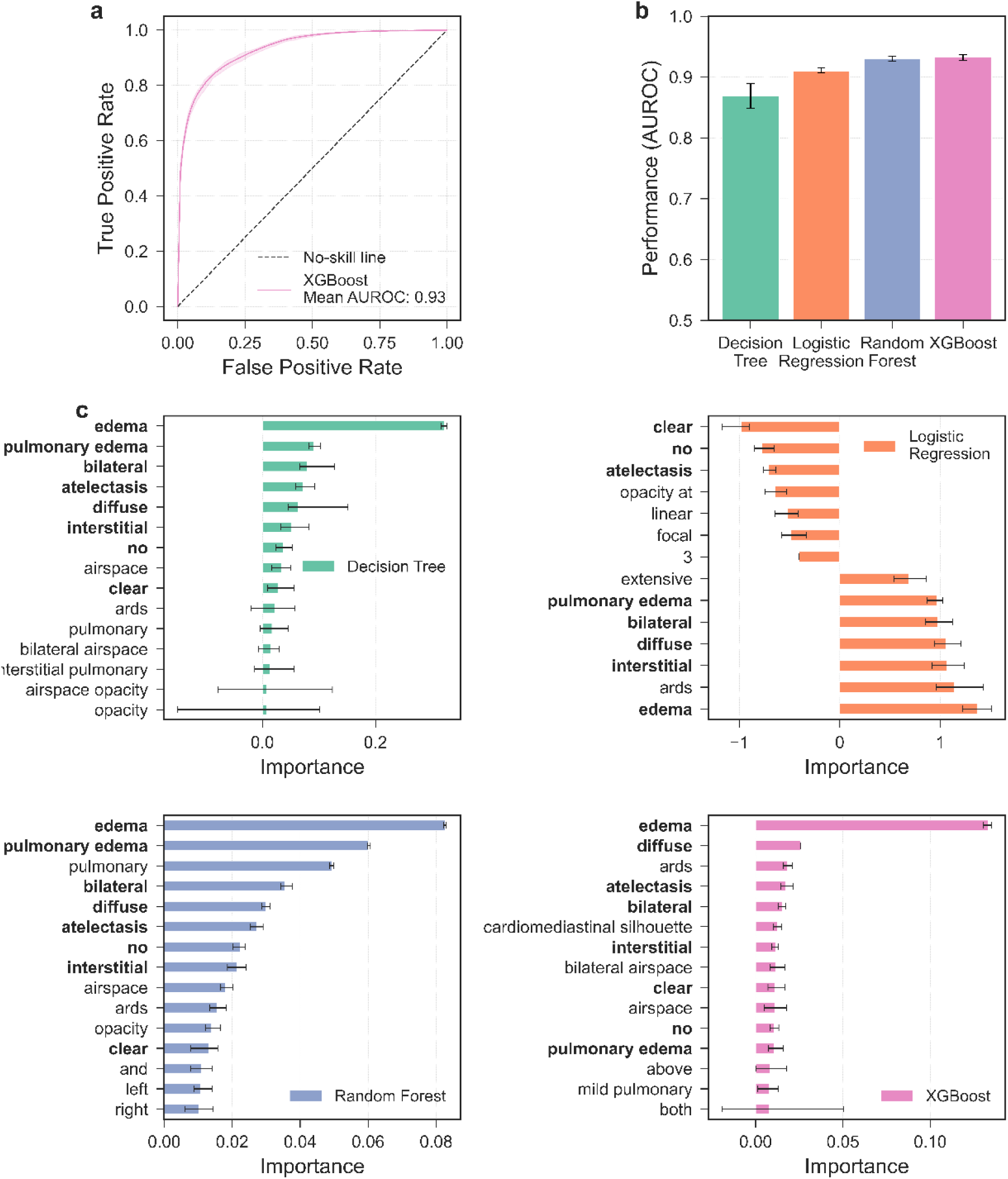
Machine learning (ML) models achieve high-performance in adjudicating the presence of bilateral infiltrates from chest imaging reports. Error bars and bands show 95% confidence intervals for estimates of the mean obtained using bootstrapping. **a)** Receiver operating characteristic (ROC) curve for the XGBoost model trained on chest imaging reports from the development set. **b)** Bootstrapped mean area under the ROC (AUROC) show that all four ML approaches yield accuracies greater or equal to 0.85. **c)** Feature importances for the four different ML approaches considered. Features in bold are highly ranked in importance in all 4 approaches.

We calculated the importance that each model assigned to the 200 tokens used as features. For decision trees and random forests, feature importance corresponds to the mean decrease in Gini impurity; for logistic regression, importances correspond to the mean value of coefficients in the fitted linear equation; and for XGBoost, the importance corresponds to the mean gain in predictive performance obtained by including a particular feature in the trees. Reassuringly, we find that the four models consistently identify tokens such as edema, bilateral, clear, and atelectasis as the most predictive (Fig. 1c). These tokens correspond closely to the inclusion/exclusion language we developed to address Berlin Definition shortcomings^8^, which we also observed when implementing Shapley-additive explanations (SHAP) values to assess feature importance when applying the Bilateral Infiltrates Model (see methods, and below) on chest imaging reports from MIMIC (2001-12) (Fig. S2). For example, the presence of the terms ‘bilateral’ or ‘edema’ has a positive impact on classification as positive for bilateral infiltrates. In contrast, the presence of the terms ‘clear’, ‘left’, or ‘right’ has a negative impact on classification as positive for bilateral infiltrates.

We then assessed how calibrated the model output probabilities were by comparing model output probabilities after training to actual rate of occurrence of positive labels in chest imaging reports from the development set. Figure 2 suggests that logistic regression and XGBoost predicted probabilities are well calibrated, which is expected given their use of similar loss functions for fitting (log-loss). In contrast, random forest produces poorly calibrated probabilities, being over-confident when forecasting with confidence levels lower than 50%, and under-confident with confidence levels greater than 50%. We thus select XGBoost as the implemented classifier for our pipeline as it offers the highest predictive performance (AUROC = 0.93, 95%CI = [0.93, 0.94]) and well-calibrated forecasts on the development set (Durbin-Watson statistic = 1.65, 95%CI = [1.35-1.92]).

**Figure 2.**
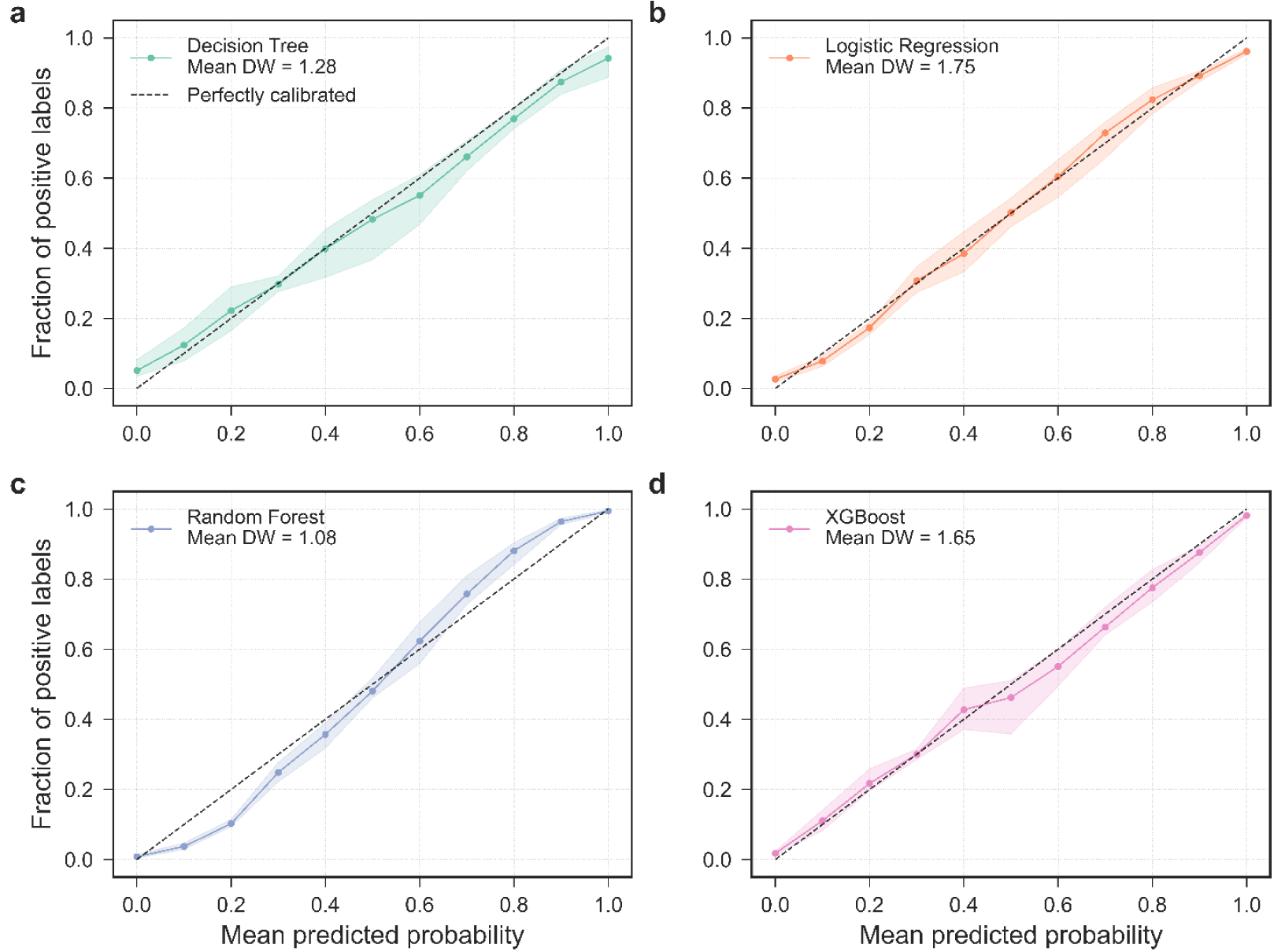
Calibration of output probabilities in implemented ML models. Error bands show 95% confidence intervals for estimates of the mean obtained using bootstrapping. We show calibration curves for models trained on the development set: **a)** decision tree, **b)** logistic regression, **c)** random forest, and **d)** XGBoost. A perfectly calibrated model would have a 1:1 relationship between the fraction of positive labels and mean probabilities (i.e., it would overlay the diagonal line). The Durbin-Watson statistic, DW, probes for correlations in the residuals, if DW is close to 2, then one can rule out correlations in the residuals, implying good linear behavior.

To assess how an XGBoost model developed for a specific cohort generalizes to a different health system dataset, we tested the discriminative performance of an XGBoost model trained on Hospital A (2013) on chest imaging reports from Hospital B (2017-18) and MIMIC (2001-12), separately (Fig. S3a). We also trained a second XGBoost model on chest imaging reports from Hospital A (2016) and tested this model against chest imaging reports from Hospital B (2017-18) and MIMIC (2001-12), separately (Fig. S3b). In general, we found both XGBoost models performing at a mean AUROC of 0.85 or higher (lowest 95%CI of 0.84, highest 95%CI of 0.93). These results show that XGBoost could stably generalize well regardless of which available hospital system we chose to use for training.

To assess generalizability, we tuned hyperparameters and trained an XGBoost model, which we name the Bilateral Infiltrates (BI) model, using the entire development set. We tested this BI model on 975 labeled chest imaging reports from MIMIC (2001-12), achieving a mean AUROC of 0.88 (95% CI: [0.85-0.90]), consistent with cohort-specific XGBoost models mentioned previously (Fig. 3a). However, calibration indicated the model was generally overconfident, except at probabilities below 10% (Fig. 3b). In addition, since prior evaluations were per report, we also assessed performance on a per-encounter basis by randomly selecting one report per encounter and applying the BI Model to those reports. Results were comparable to the per-report analysis, though with a wider 95% confidence interval due to the smaller sample size (100 vs. 975 reports, Fig. S4).

**Figure 3.**
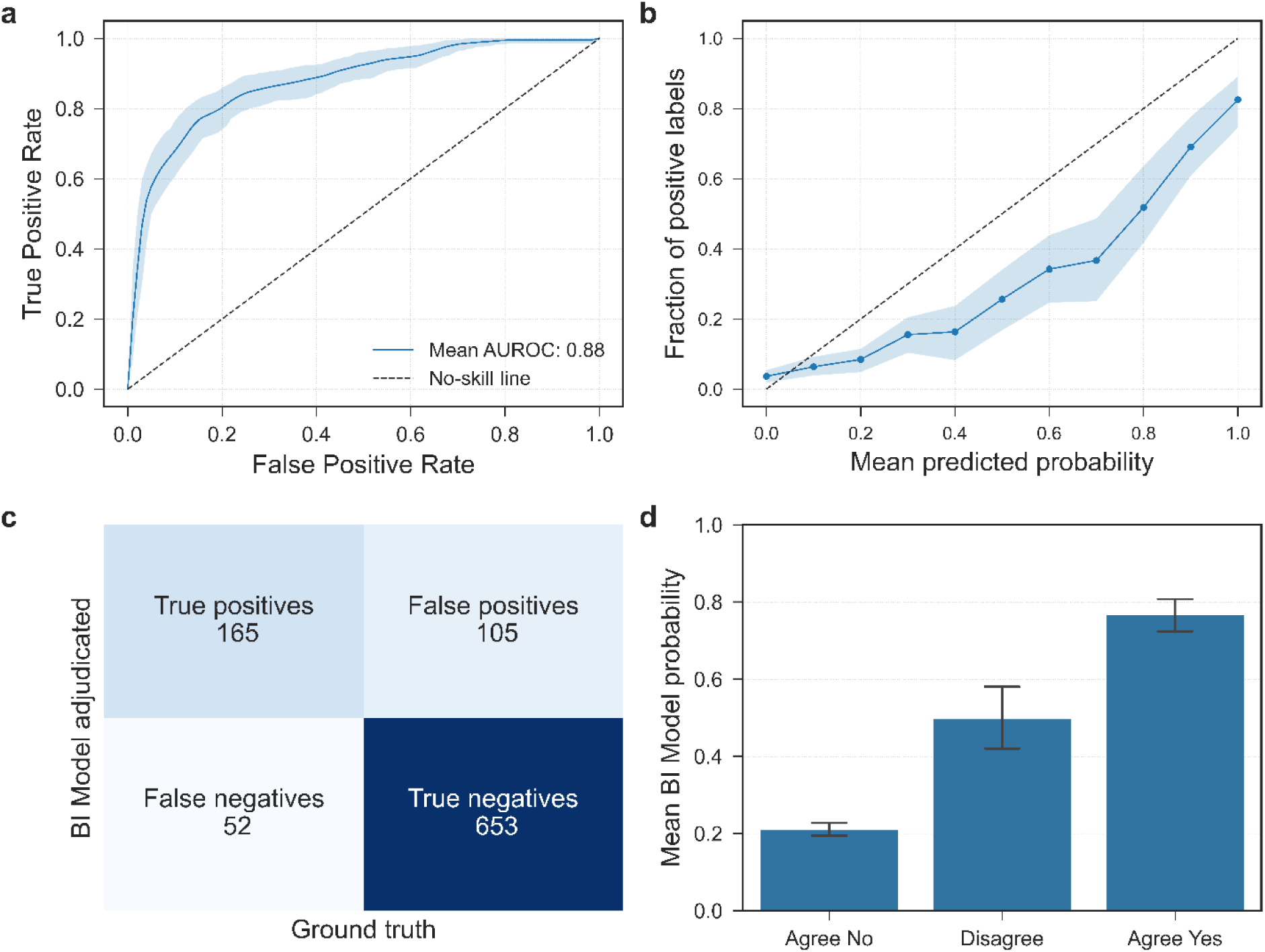
Evaluation of the Bilateral Infiltrates (BI) model on chest imaging reports from MIMIC (2001-12). Error bars and bands show 95% confidence intervals for estimates of the mean obtained using bootstrapping. **a)** ROC curve for the BI model tested on 975 bootstrapped chest imaging reports from MIMIC (2001-12). **b)** Calibration of probabilities by the BI model when applied on 975 bootstrapped chest imaging reports from MIMIC (2001-12). **c)** Confusion matrix comparing MIMIC (2001-12) chest imaging report adjudications by the critical care physician (ground truth) against BI model adjudications done at a 50% probability threshold. Notice that the numbers add up to 975. **d)** Comparing the output probabilities of the BI model (a measure of its confidence that a report is consistent with bilateral infiltrates) in three agreement scenarios between the critical care physician and the internal medicine physician when adjudicating MIMIC (2001-12)’s chest imaging reports.

The calibration results prompted us to test whether the output probabilities by the BI model could be associated with the inter-rater disagreement rates of the test set. To this end, we grouped MIMIC (2001-12) chest imaging reports into three groups: reports where both physician raters agreed on the ‘No’ label, reports where both physician raters agreed on the ‘Yes’ label, and reports where physician raters disagreed. For each of these groups, we calculated the mean BI Model output probability. As seen on Figure 3d, we reassuringly find that: (1) when physicians agree that a report should be adjudicated as ‘No’ for bilateral infiltrates, the BI model typically outputs 21.1% (95%CI = [19.4%, 22.7%]); (2) when physicians agree that a report should be adjudicated as ‘Yes’ for bilateral infiltrates, the BI model typically outputs 76.6% (95%CI = [72.5%, 80.8%]); (3) when physicians disagree on their bilateral infiltrate adjudication, the BI Model typically outputs 49.8% (95%CI = [41.9%, 57.7%]). This suggests that despite the BI model yielding uncalibrated probabilities for the test set, these probabilities still strongly align with expectation.

Finally, we evaluated how the BI model fared adjudicating bilateral infiltrates on individual chest imaging reports. We observed that, at the 50% probability threshold for binarizing probabilities into yes/no decisions, the BI model exhibited a 76.0% sensitivity (24.0% false negative, or missed, rate), a 13.9% false positive rate, a precision or positive predictive value of 61.1%, and a 92.6% negative predictive value in adjudicating bilateral infiltrates at the single report level (Fig. 3c). This results in an F_1_ score of 0.678 and an accuracy of 0.839. We also tried a sweep of different probability cutoffs to find which threshold maximizes the above metrics for the BI model. These can be found on Table S1, and we observe that no single probability cutoff optimizes accuracy and F_1_ score simultaneously.

### Extracting ARDS risk factors in attending physician notes

We first developed regex patterns to match keywords for risk factors and heart failure language (see SI Regular expression list 1 for a full list of regular expressions). As shown in Figure S5, most of the developed regex patterns capture 100% of the notes that were labeled ‘yes’ for a particular risk factor in Hospital A (2013). When we count the total number of notes labeled as either yes or no, the most prevalent matches were sepsis (744 notes labeled vs. 748 notes regex-captured), pneumonia (636 notes labeled vs. 955 regex-captured), and shock (604 notes labeled vs. 607 regex-captured). For the heart failure criteria, the relevant matches were the ‘cardiogenic’ keyword (176 notes labeled vs. 725 regex-captured) to qualify the matching of shock, and congestive heart failure (254 notes labeled vs. 352 regex-captured). We thus feel confident that the built regex patterns can match nearly the entirety of Hospital A (2013) notes labeled as “yes” for specific risk factors.

Next, we trained separate XGBoost models on attending physician notes from Hospital A (2013) to adjudicate pneumonia, aspiration, congestive heart failure, and sepsis. We attempted an ML approach for these because at least 100 attending physician notes were labeled for them, and their labels have relatively balanced yes/no proportions after regex-matching (between 33% and 66%, except for sepsis; see Table S2). We observed that the XGBoost model trained for pneumonia yielded the best discriminative performance during cross-validation (Pneumonia: AUROC = 0.92, 95%CI = [0.89, 0.94]; CHF: AUROC = 0.77, 95%CI = [0.58, 0.85]; Aspiration: AUROC = 0.61, 95%CI = [0.45, 0.78]; Sepsis: AUROC = 0.71, 95%CI = [0.58, 0.82]; Fig. 4a and 4b), the best calibration (DW = 1.70, 95%CI = [1.13-2.22]; Fig. 5b), assigned the greatest and most stable importance to tokens that align with clinical expectations (Fig. 4c), and exhibited the best calibration (DW = 1.72, 95%CI = [1.31-2.28]; CHF: DW = 1.21, 95%CI = [0.41, 1.85]; Aspiration: DW = 0.61, 95%CI = [0.09, 1.35]; Sepsis: DW = 0.90, 95%CI = [0.30, 1.68]; Fig. 5d). These results reveal a stark contrast between the XGBoost models trained to adjudicate pneumonia and the XGBoost models trained to adjudicate congestive heart failure, aspiration, and sepsis. Thus, we decided against integrating these three ML models into our pipeline and only use the XGBoost approach to adjudicate pneumonia.

**Figure 4.**
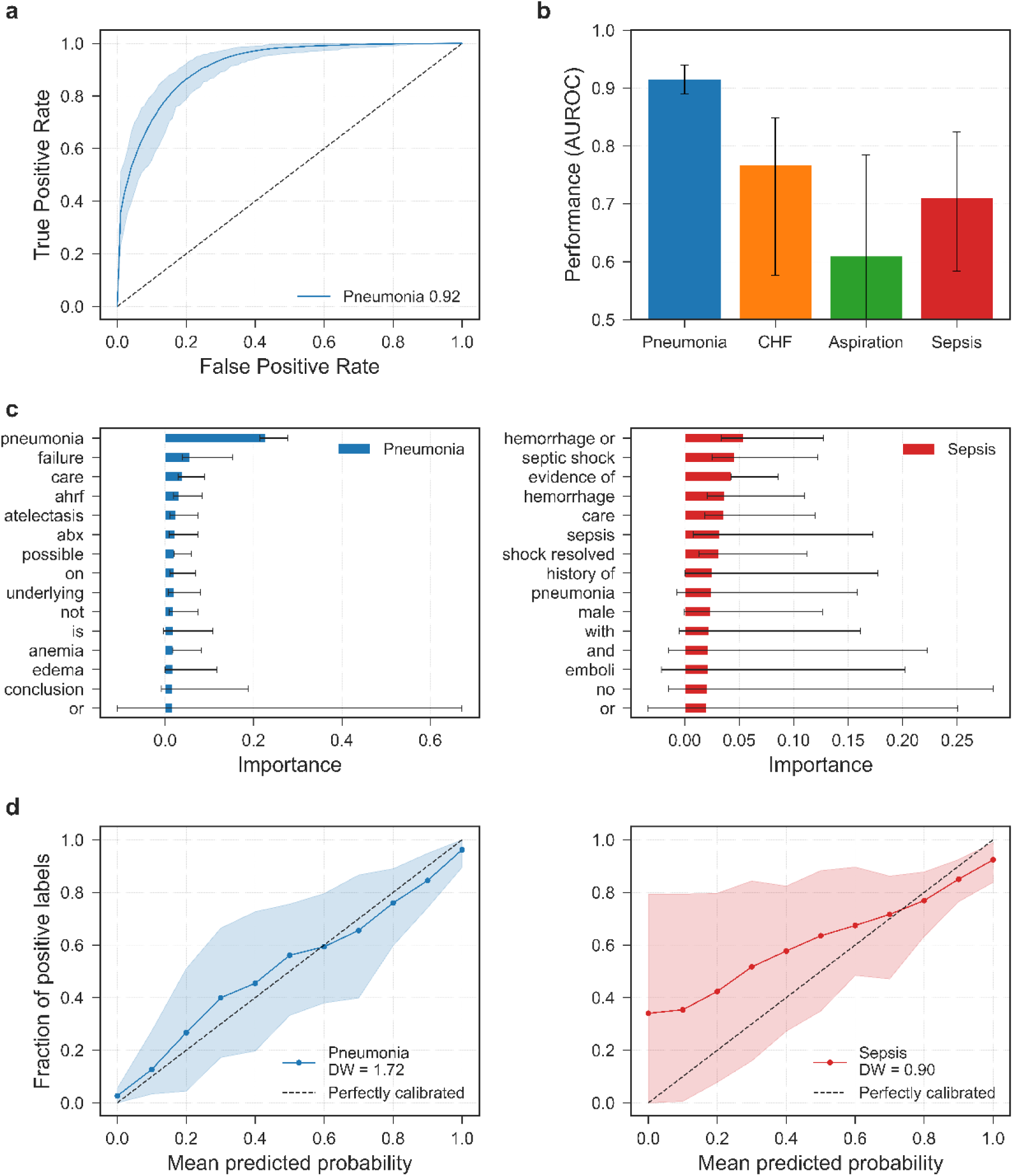
XGBoost model performance in adjudicating the presence of some risk factors in attending physician notes amenable to ML techniques. Error bars and bands show 95% confidence intervals for estimates of the mean obtained using bootstrapping. **a)** ROC curve showing the cross-validated performance of XGBoost model trained to adjudicate pneumonia on Hospital A (2013) attending physician notes. **b)** Bar plot showing cross-validated AUROCs of XGBoost models trained to adjudicate all the attempted risk factors, plus congestive heart failure, on Hospital A (2013) attending notes. **c)** Training set feature importances by the XGBoost models trained to adjudicate pneumonia and sepsis. **d)** Training set calibration curves for the XGBoost models trained to adjudicate pneumonia and sepsis.

**Figure 5.**
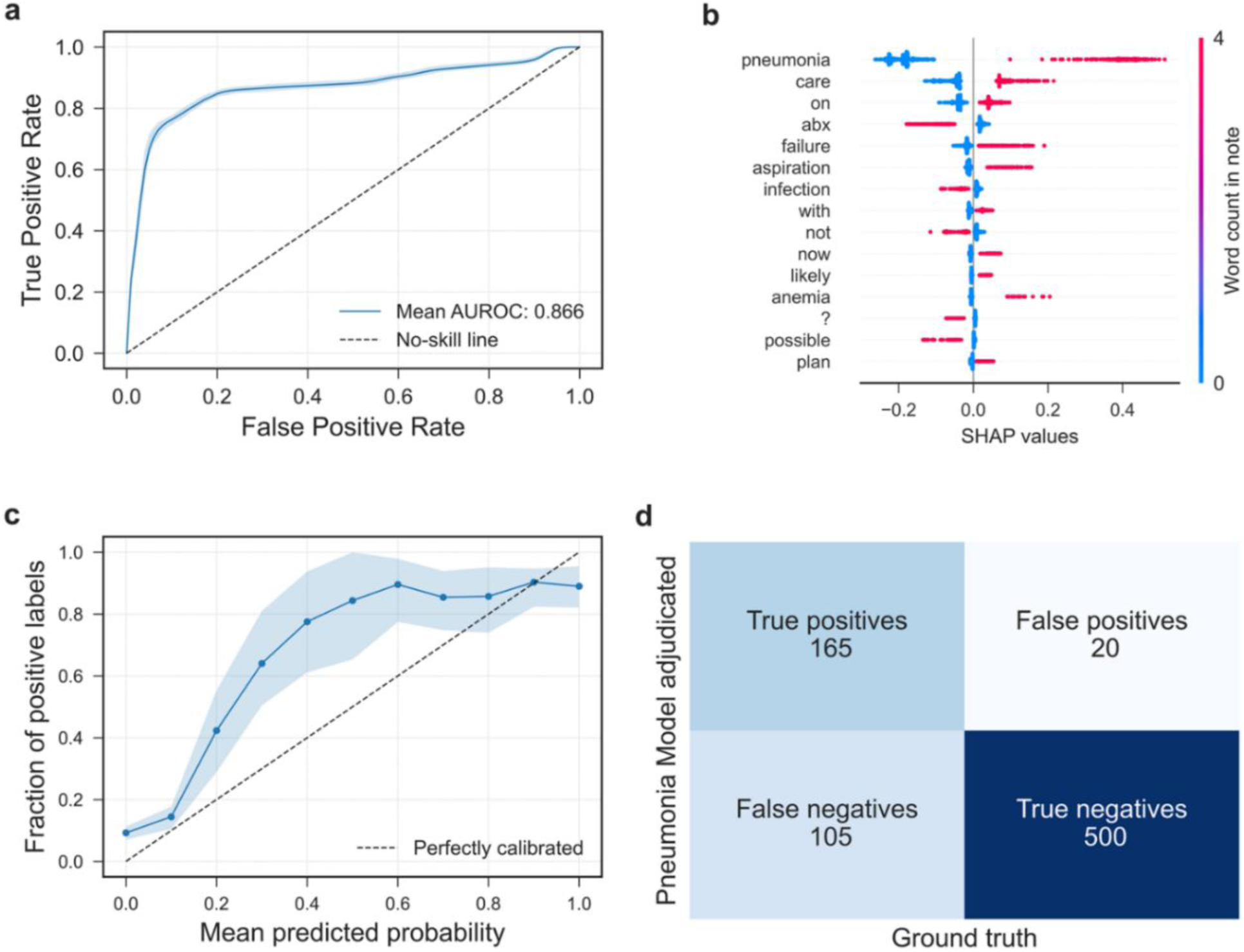
Evaluation of the Pneumonia Model on attending physician notes from MIMIC (2001-12). Error bands show 95% confidence intervals for estimates of the mean obtained using bootstrapping. **a)** ROC curve for the Pneumonia Model tested on 790 bootstrapped attending physician notes from MIMIC (2001-12) that were regex-captured for pneumonia. **b)** SHAP values for the top 15 words in terms of their impact on Pneumonia Model’s output probabilities. **c)** Calibration of probabilities by the Pneumonia Model when applied on 790 bootstrapped attending physician notes from MIMIC (2001-12). **d)** Confusion matrix comparing MIMIC (2001-12) attending physician notes pneumonia labels by the critical care physician (ground truth) against Pneumonia Model adjudications done at a 50% probability threshold. Notice that the numbers add up to 790.

We then evaluated our approach for adjudicating pneumonia by training an XGBoost classifier on 955 attending physician notes regex-captured for pneumonia. The resulting Pneumonia Model was tested on 790 regex-captured notes from MIMIC (2001-12), achieving a mean AUROC of 0.87 (95%CI = [0.84-0.89]) (Fig. 5a). The model relied heavily on the ‘pneumonia’ token for classification (Fig. 5b). Calibration analysis indicated general under-confidence, except at probabilities above 80% (Fig. 5c), in contrast to the overconfidence observed in the Bilateral Infiltrates model.

Finally, we evaluated the Pneumonia Model’s performance on individual attending physician notes. At a 50% probability threshold, it achieved 61.1% sensitivity (38.9% false negative rate), 3.8% false positive rate, 89.2% precision, and 82.6% negative predictive value, yielding an F_1_ score of 0.725 and an accuracy of 0.842 (Fig. 5d). We also explored various probability cutoffs to optimize these metrics (Table S3). Notably, the narrow range 19.3%–19.8% emerged as optimal for both the F_1_ score and Youden’s J statistic.

For other risk factors and heart failure criteria, we used regex adjudication via two approaches. In the first, if a factor was labeled ‘yes’ in >80% of captured notes, we adjudicated it using the same regex pattern, including shock, cardiogenic, inhalation, pulmonary contusion, vasculitis, drowning, and overdose (Table S2 and Fig. S6). In the second, to reduce false positives, we applied exclusion terms after regex capture (Regular expression list 2). This method, used for sepsis and shock, significantly lowered false positive rates compared to regex alone (Fig. S7).

We then applied these regex approaches to adjudicate risk factors or heart failure criteria in MIMIC (2001-12) attending physician notes, focusing on factors with available labels to validate patterns developed for Hospital A (2013). These regex patterns demonstrated strong performance, particularly for burns, pancreatitis, sepsis, and aspiration (Fig. 6).

**Figure 6.**
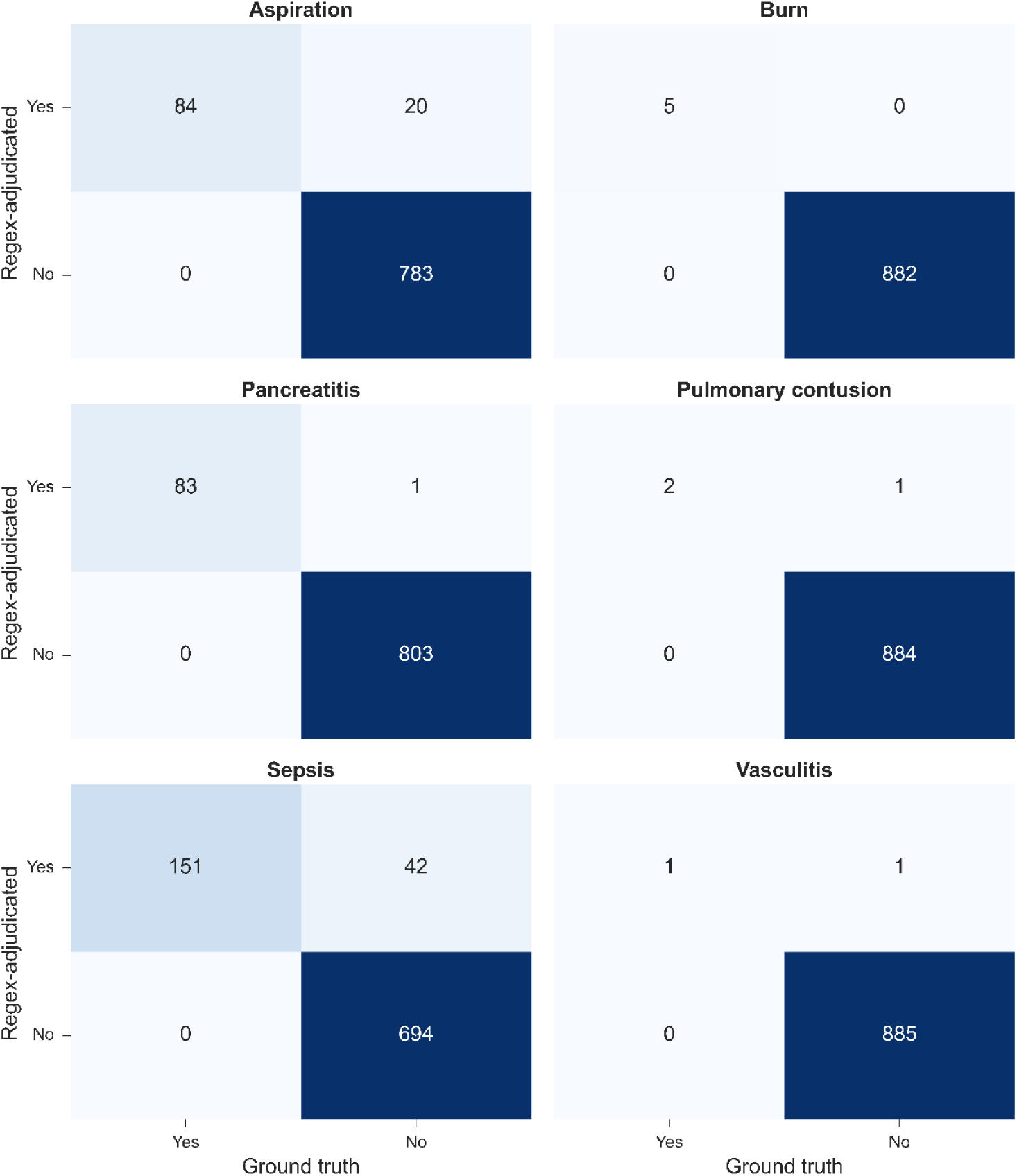
Risk factor adjudication performance of regular expressions on MIMIC (2001-12). All regular expressions developed adjudicate 100% of the attending physician notes from MIMIC (2001-12) labeled as “yes” for each risk factor. In terms of precision, burns, pancreatitis, sepsis, and aspiration exceeded 80%.

Based on the overall results, we incorporated regex patterns to the pipeline to adjudicate the following risk factors or heart failure language: *pneumonia, sepsis, shock and its “cardiogenic” qualifier, aspiration, inhalation, pulmonary contusion, pancreatitis, burns, vasculitis, drowning, and overdose*.

### Adjudication of heart failure from echocardiogram (echo) reports

The criteria for the objective assessment of heart failure rely on the following six factors: left ventricular ejection fraction, cardiopulmonary bypass, left atrial diameter, left atrial volume index, left ventricular hypertrophy, and grade II or III diastolic dysfunction. Because echo reports are highly standardized, it is possible to extract these factors from the reports using regex. Moreover, we had access to echo reports from Hospital A (2013) which were previously text-matched, enabling us to validate our regex approach.

Using the regex patterns listed in the SI, we analyze Hospital A (2013)’s echo reports for the presence or absence of each of the six factors of interest. Figure 7 demonstrates that not all six factors were present in every echo report. For three of the six factors — left ventricular ejection fraction, left atrial dimension/diameter, and left atrial volume index— we found excellent agreement between regex and text-matching. Two of the other three, cardiopulmonary bypass and diastolic function, were not text-matched, so no comparison can be made. For left ventricular hypertrophy, the regex-matching procedure correctly captured the desired language, indicating that the original text-matching procedure failed to identify 13 echo reports. In addition, we validated the numerical values extracted through this regex approach by randomly selecting 10% of echo reports for visual inspection of values and comparing against values extracted through regex. We found 100% concordance between values extracted and those retrieved manually (Table S4).

**Figure 7.**
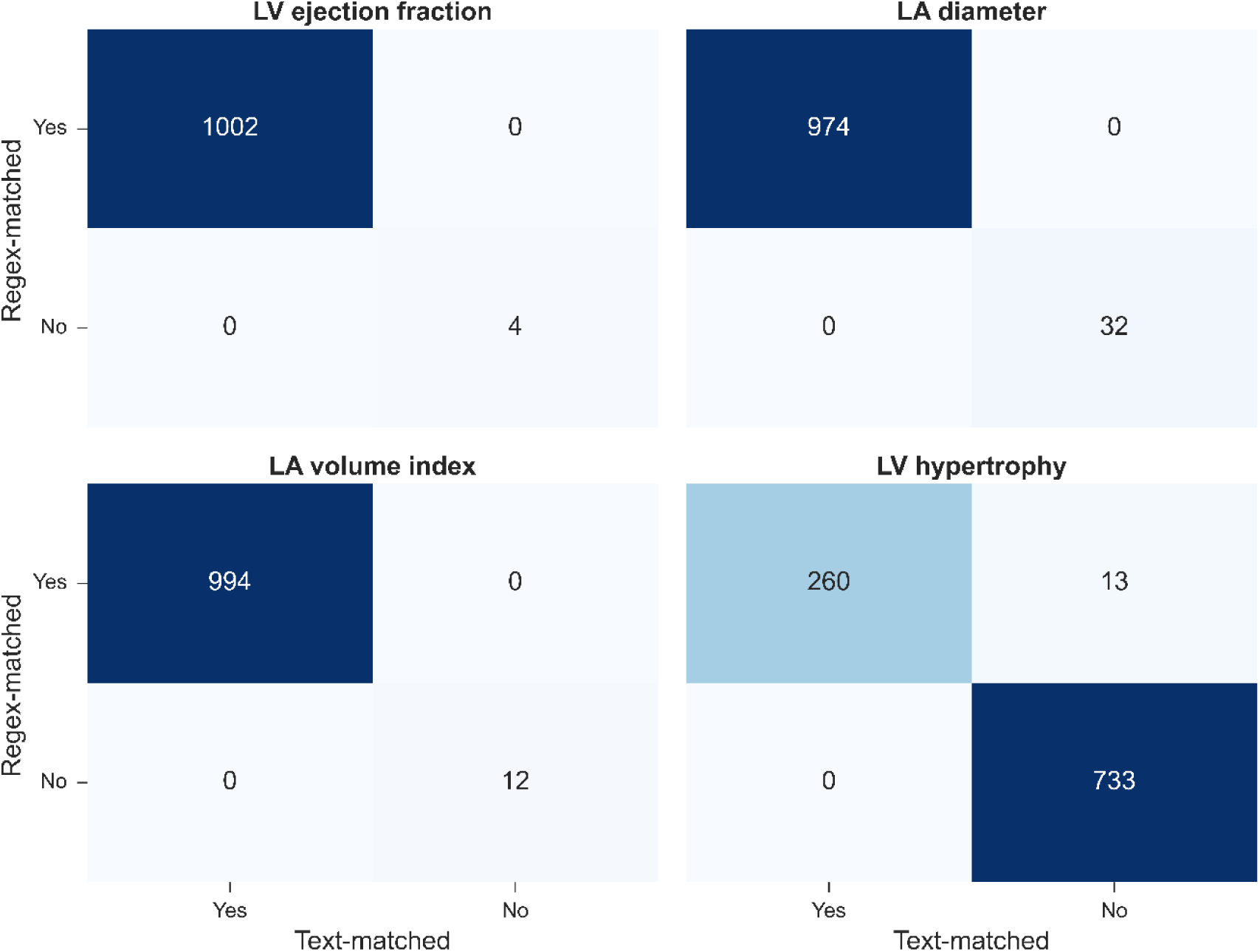
Confusion matrices comparing the flagging performance regex-matching against text-matching for. Left ventricular ejection fraction, Left atrial dimension/diameter, Left atrial volume index, and Left ventricular hypertrophy on echo reports from Hospital A (2013) Note the large discrepancy for the annotations of ‘left ventricular hypertrophy’, which is explained in text.

### Adjudication of ARDS for Hospital A (2013) cohort

We are now ready to compare the performance of our complete pipeline against the previously reported ARDS adjudication^8^. We use a threshold of 50% to map output probabilities into binary yes/no decisions for both the Bilateral Infiltrates model and the Pneumonia Model.

We conduct the evaluation of our pipeline for the 943 encounters in the Hospital A (2013) ARDS adjudication cohort who were 18 years and older, received invasive mechanical ventilation, and had acute hypoxemic respiratory failure (Fig. 8). 143 encounters had no chest imaging report available and were adjudicated as negative for ARDS. The remaining 800 encounters had at least one chest imaging report available. The BI model adjudicated bilateral infiltrates within 48 h of a hypoxemic episode for 510 encounters. Of these 510 encounters, 443 had at least one of the qualified hypoxemic events occurring post-intubation and 333 had a risk factor within 7 days of the qualified hypoxemia event and were adjudicated as being positive for ARDS.

**Figure 8.**
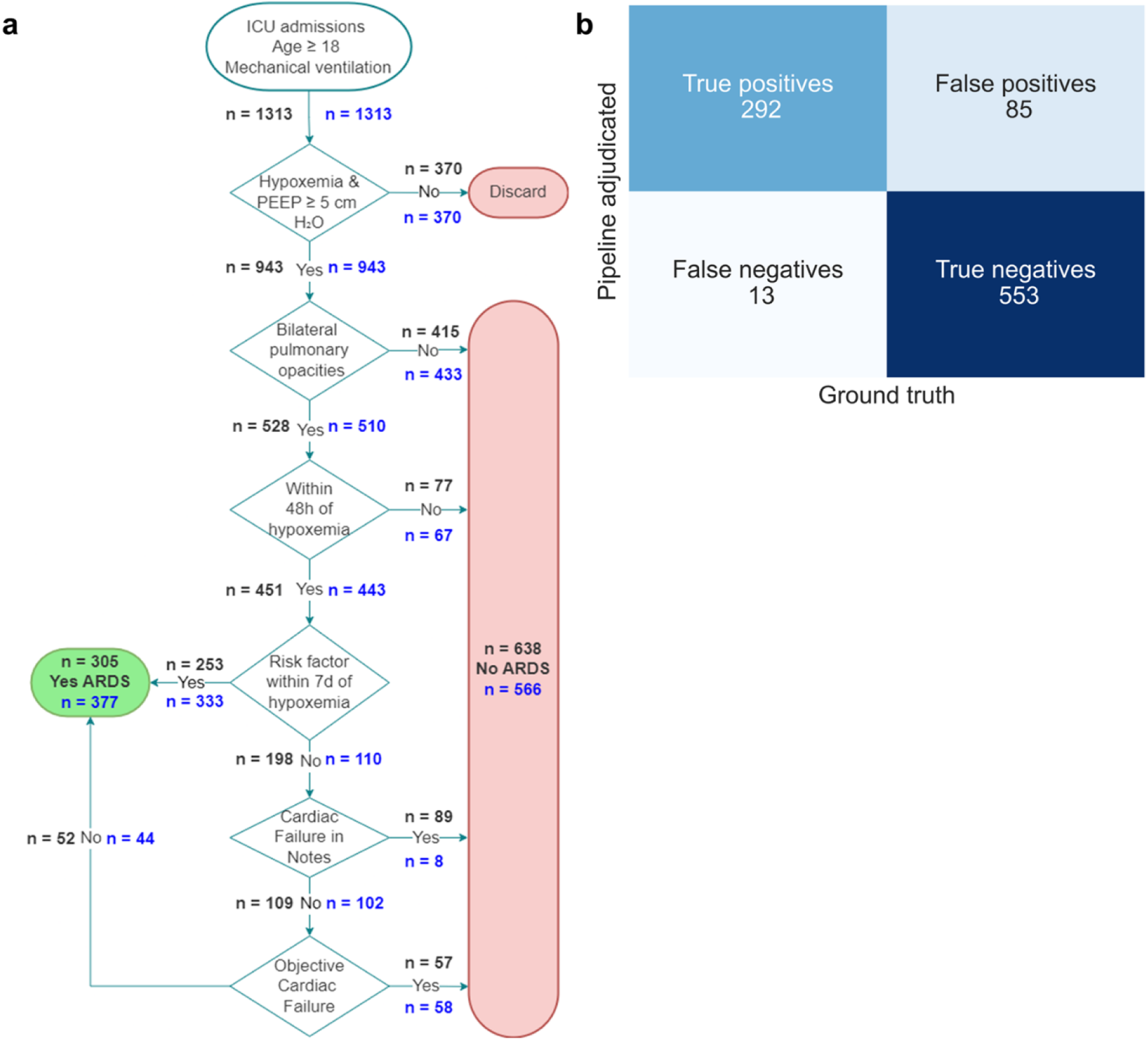
Computational pipeline for adjudication of Hospital A (2013) cohort yields a small fraction of false negatives and an acceptable fraction of false positives. **a)** Flowchart of ARDS adjudication by computational pipeline (blue) vs. physician (black). **b)** Confusion matrix comparing physician adjudication from previous publication^8^ against computational adjudication pipeline.

The remaining 110 encounters were then evaluated for heart failure. For 8 encounters, the physician notes indicated heart failure, and they were adjudicated as negative for ARDS. The last 102 encounters were then adjudicated using the objective heart failure assessment step; 58 were adjudicated to have heart failure and thus negative for ARDS and the remaining 44 were adjudicated as positive for ARDS. In total, the pipeline adjudicated 377 encounters as positive for ARDS and 566 as negative for ARDS.

To summarize, using a standard 50% probability cutoff for both ML models, our pipeline yields close agreement with the physician adjudication of ARDS for this cohort^8^ (Fig. 8a). Specifically, the pipeline yields a sensitivity or true positive rate of 95.7% on this cohort, which compares most favorably to the 12.2% ARDS documentation rate we found on this cohort^23^. Importantly, this high sensitivity is achieved while maintaining a low 13.3% rate of false positives.

As a sensitivity analysis, we explored different probability thresholds for the BI model to identify optimal metrics for both the model and the overall adjudication pipeline (Table S5). A BI model cutoff between 57.4% and 58.4% yielded the best overall accuracy (0.902), F_1_ score (0.864), and Youden’s J statistic (0.834), while also optimizing the false positive rate (12.4%) and precision (78.7%). However, thresholds optimizing the BI model’s F_1_ score (27.1%–29.2%) resulted in a lower false negative rate (2.9%) and higher negative predictive value (98.3%) for the overall pipeline.

Finally, we compared the clinical characteristics of encounters adjudicated as ARDS-positive or negative by both the pipeline and critical care physician adjudication^8^ (Table 2). Both consistently identified a clinically distinct population. Encounters adjudicated with ARDS had lower PF ratios, higher rates of low tidal volume ventilation, and worse mortality and length of stay outcomes.

**Table 2.** Clinical characteristics of Hospital A (2013) encounters adjudicated as yes/no ARDS by the pipeline and a critical care physician.

| Characteristic | ARDS |  | Not ARDS |  |
| --- | --- | --- | --- | --- |
|  | Pipeline | Intensivist | Pipeline | Intensivist |
| Age (years), median [IQR] | 62<br>[51-71] | 62<br>[50-70] | 63<br>[52-73] | 63<br>[52-73] |
| PF ratio (mm Hg), median [IQR] | 201.3 [132-280] | 207.5 [140-286.7] | 272.5 [177.5-355.8] | 242.9 [152.5-340] |
| Encounters that received Low Tidal Volume Ventilation at any time, n/N (%) | 74/370 (20.0%) | 63/302 (20.9%) | 23/743 (3.1%) | 34/811 (4.2%) |
| Encounters with plateau pressure > 30 cm H2O at any time, n/N (%) | 63/366 (17.2%) | 55/301 (18.3%) | 17/687 (2.5%) | 25/752 (3.3%) |
| In-hospital mortality, n/N (%) | 143/377 (37.9%) | 120/305 (39.3%) | 83/852 (9.7%) | 106/924 (11.5%) |
| ICU Length of Stay (days), median [IQR] | 9<br>[4-17] | 9<br>[5-17] | 2<br>[1-4] | 2<br>[1-4] |

### Adjudication of ARDS for MIMIC (2001-12) labeled subset

We applied our automated ARDS adjudication to 100 randomly selected balanced encounters in MIMIC (2001-12) and then compared a critical care physician’s adjudication against the pipeline’s (Fig. 9). As for Hospital A (2013), we used a 50% probability threshold for the ML models within the pipeline. Reassuringly, we find that the overall performance of our pipeline on MIMIC (2001-12) is strikingly similar to its performance on the Hospital A (2013) cohort (Fig. 9b). Specifically, the pipeline yields a sensitivity or true positive rate of 93.5% on MIMIC (2001-12), which compares favorably to the 22.6% ARDS documentation rate we found in this subset (see SI for discussion relating to a classification difference for mild ARDS in this cohort). This high sensitivity is achieved while maintaining a relatively low 17.4% false positive rate. Moreover, the false negative rate of the pipeline adjudication (6.5%) is lower than that of a physician not trained in critical care medicine (16.1%), highlighting the pipeline’s potential to aid physicians recognize ARDS.

**Figure 9.**
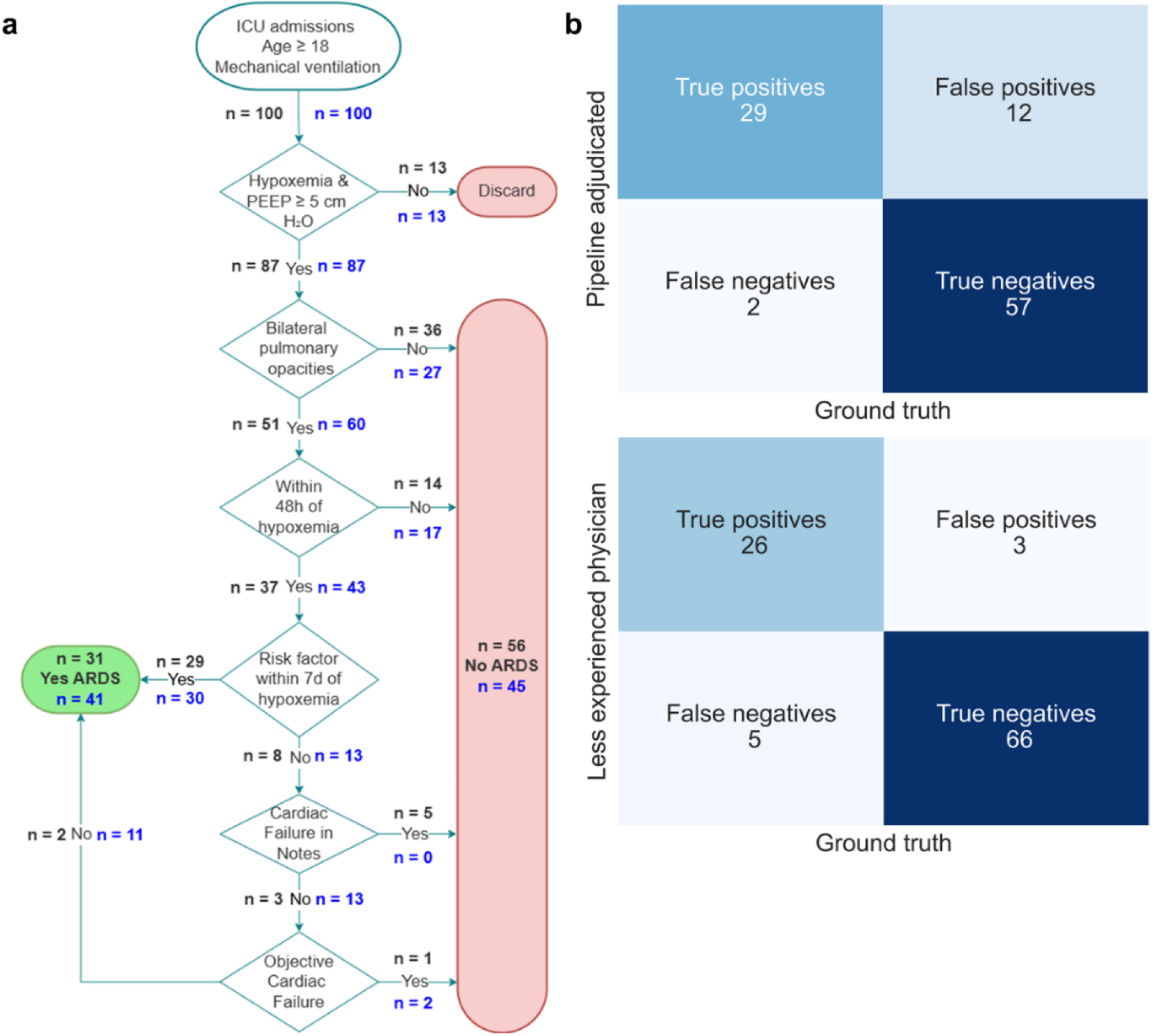
Computational pipeline for adjudication of MIMIC (2001-12) cohort yields a small fraction of false negatives and a manageable fraction of false positives. **a)** Flowchart of ARDS adjudication by computational pipeline (blue) vs. physician (black). **b)** Confusion matrix comparing physician adjudication (ground truth) against computational adjudication pipeline (top panel), and physician adjudication (ground truth) against a less experienced physician adjudication.

We also explored probability thresholds for the BI and Pneumonia Models to identify optimal metrics for each model and the overall pipeline (Table S6). A BI Model cutoff of 84.7% and 86.6% and a Pneumonia Model cutoff of 16.0% to 80.7% yielded the best overall accuracy (0.890), F_1_ score (0.831), and Youden’s J statistic (0.770). Like Hospital A (2013), this range also optimized the false positive rate (10.1%) and precision (79.4%). However, thresholds maximizing each model’s F_1_ score individually resulted in a lower false negative rate (6.5%) and higher negative predictive value (96.6%) for the pipeline.

Finally, we assessed the clinical characteristics of encounters classified as ARDS-positive or negative by both the pipeline and the critical care physician (Table 3). As seen in Hospital A (2013), both methods identified a clinically distinct population. ARDS-positive encounters had lower PF ratios, a higher proportion receiving low tidal volume ventilation, and longer hospital stays. However, mortality rates were similar, likely due to the small sample size (100 encounters) and only 11 deaths. We also found that diagnosis occurred within 48 hours of intubation for most encounters (see the SI for details).

**Table 3.**
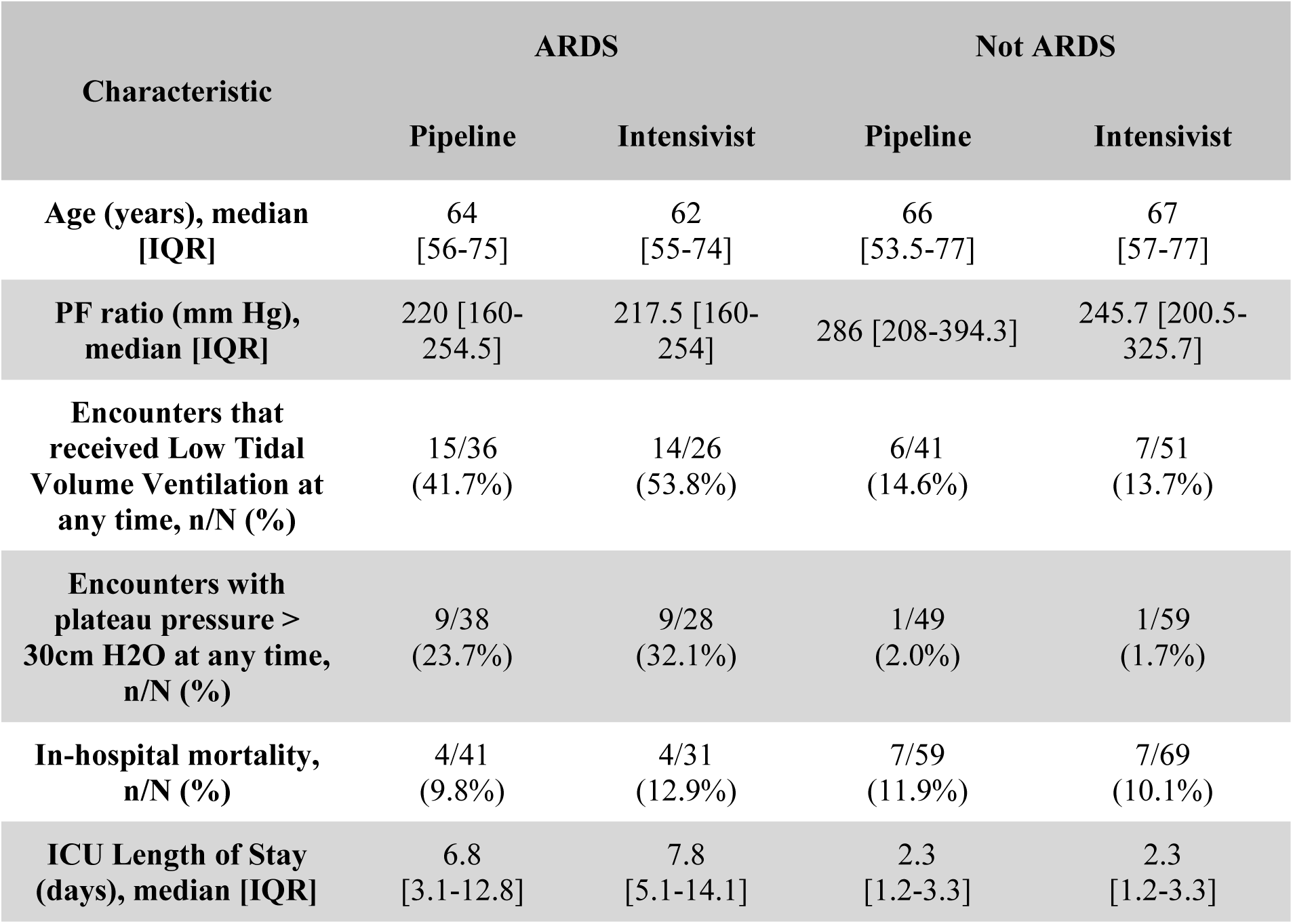
Clinical characteristics of MIMIC (2001-12) encounters adjudicated as yes/no ARDS by the pipeline and a critical care physician.

| Characteristic | ARDS |  | Not ARDS |  |
| --- | --- | --- | --- | --- |
|  | Pipeline | Intensivist | Pipeline | Intensivist |
| Age (years), median [IQR] | 64 [56-75] | 62 [55-74] | 66 [53.5-77] | 67 [57-77] |
| PF ratio (mm Hg), median [IQR] | 220 [160-254.5] | 217.5 [160-254] | 286 [208-394.3] | 245.7 [200.5-325.7] |
| Encounters that received Low Tidal Volume Ventilation at any time, n/N (%) | 15/36 (41.7%) | 14/26 (53.8%) | 6/41 (14.6%) | 7/51 (13.7%) |
| Encounters with plateau pressure > 30cm H2O at any time, n/N (%) | 9/38 (23.7%) | 9/28 (32.1%) | 1/49 (2.0%) | 1/59 (1.7%) |
| In-hospital mortality, n/N (%) | 4/41 (9.8%) | 4/31 (12.9%) | 7/59 (11.9%) | 7/69 (10.1%) |
| ICU Length of Stay (days), median [IQR] | 6.8 [3.1-12.8] | 7.8 [5.1-14.1] | 2.3 [1.2-3.3] | 2.3 [1.2-3.3] |

## Discussion

In this study, we developed and validated a computational pipeline to automate ARDS adjudication per the Berlin Definition using EHR data. Our approach combined high-performing XGBoost models for analyzing chest imaging reports and physician notes with regular expressions and structured data to assess ARDS risk factors and rule out heart failure. The pipeline achieved excellent test characteristics on Hospital A (2013), with false negative and false positive rates of 4.3% and 12.4%, respectively, at the optimal probability cutoff for accuracy and F_1_ score. Validation on a MIMIC-III subset showed generalizability, yielding false negative and false positive rates of 12.9% and 10.1%, respectively, at optimal probability cutoffs for accuracy and F_1_ score.

A question that readers may ask is why we pursued this approach instead of using increasingly popular large language models (LLMs). We believe that in this and many other health related contexts, using an interpretable model that can be easily versioned offers benefits that would be lost even if an LLM could hypothetically produce higher accuracy. Specifically, we believe that computational pipelines aiding complex diagnoses should follow two principles. First, they should assist, not replace, physicians—flagging potential diagnoses rather than mandating them. Second, they should provide interpretable insights. Others have pointed out^24^ that machine learning (ML) should only serve as a final decision-maker for deterministic tasks (e.g., distinguishing a dog from a cat). In contrast, for tasks with overlapping class characteristics and significant consequences, such as medical diagnosis, ML should estimate probabilities rather than make definitive decisions. Consequently, the tree-based methods we consider estimate probabilities, the first stage of any classification problem, and enable physicians to optimize the false positive vs. false negative tradeoff by adjusting the probability cutoff. In addition, our XGBoost implementation allows language-level interpretation of probabilities, which can enhance physician trust in ML models.

We present our pipeline here as a tool to assist healthcare quality reviewers during retrospective reviews. Its greatest potential, however, lies in clinical decision support, providing timely ARDS alerts to critical care physicians. Importantly, any ‘live’ implementation must consider the precepts of decision theory, as our XGBoost models generate probabilities that require binarization into “yes” or “no” based on a chosen threshold balancing false positives and negatives. Our perspective is that given ARDS’s under-recognition, prioritizing low false negative rates is crucial for life-saving interventions like low tidal volume ventilation or prone positioning^25^. However, over-recognition carries risks, such as unnecessary treatments (e.g. placing a non-ARDS patient in prone position or treating a non-ARDS patient with low tidal volume ventilation or neuromuscular blockade^26,27^) or alert fatigue^28–30^. Nevertheless, each provider or health system should determine its own threshold according to their own preferences or policies, guided by decision theory frameworks like Youden’s J statistic (for equal weighting of false positives and negatives)^31,32^, or the Net Benefit method (which uses the probability cutoff as a false positive to true positive ratio to assess clinical utility)^33,34^. Future studies on critical care physicians’ perspectives on this tradeoff could further inform implementation.

Our study advances efforts to automate ARDS diagnosis in key ways. First, it demonstrates automation of ARDS adjudication for intubated adult ICU patients. Second, it demonstrates that integrating open-source ML and rules-based methods enhances applicability across cohorts. Third, it demonstrates how the inclusion of multi-center and multi-timepoint data to develop ML approaches increases model robustness and generality for adjudicating chest imaging reports. Fourth, it provides a benchmark making use of a publicly available dataset, thus enabling future comparisons of performance across pipelines.

The latter advance is particularly critical since prior studies relied on single-center EHR data or non-reproducible methods. For example, Afshar et. al. used text features in chest imaging reports for ARDS identification reporting an AUROC of 0.80^35^. However, our work identifies ARDS by considering data beyond chest imaging reports. Sathe et. al. developed EHR-Berlin, evaluating the Berlin Definition using ML and rules-based methods, but their focus was limited to COVID-19 patients^16^; by using a cohort of patients who were already defined as having an ARDS risk factor, they effectively eschewed the need to identify ARDS risk factors or heart failure. In contrast, our pipeline evaluates all Berlin Definition components for mechanically ventilated adult patients. Song and Li developed a fully rules-based Berlin Definition tool, both achieving identical high performance^14,15^. However, their methods were constructed within a single hospital over a single period, which limits generalizability.

Machine learning enables efficient analysis large volumes of data that would otherwise require extensive human effort. As an example, a recent study that combined NLP techniques with manual chart abstraction reduced retrospective review time from 2,000 hours to just 34.3 hours^36^. Similarly, our pipeline adjudicates ARDS for hundreds of encounters in under five minutes by training XGBoost models at runtime, with even faster inference using pre-trained models. This automation can help mitigate ARDS under-recognition in clinical practice by consistently applying the Berlin Definition diagnostic pipeline, reducing reliance on limited human cognitive resources, especially in the demanding environment of critical care^1^.

## Limitations

Our computational pipeline achieved >90% sensitivity in identifying ARDS in Hospital A (2013) and MIMIC (2001–12) cohorts. However, this alone does not reflect its clinical impact without comparison to clinician recognition rates. We benchmarked our pipeline against ARDS documentation rates by treating physicians, though our prior work suggests documentation may underestimate true recognition^23^. Clinicians may recognize ARDS without recording it in notes or applying specific treatments. Notably, even the highest clinician recognition rate—78.5% for severe ARDS in LUNG SAFE^7^—remains below our pipeline’s >90% sensitivity, underscoring its potential to enhance ARDS recognition in clinical practice.

A limitation of our approach is its focus on intubated patients, potentially reducing applicability to mild ARDS cases, where non-invasive ventilation is now included under the updated Berlin Definition. The pipeline excludes PF ratios unless confirmed post-intubation, as it was developed before the 2024 Berlin Definition update, which expanded inclusion to patients on high-flow oxygen^37^. However, our pipeline could adapt to include these patients if nasal cannula flow rates and pulse oximetry values are available, despite challenges with non-invasive FiO_2_ delivery^38^. Future work will expand inclusion criteria accordingly.

Another limitation is the variability in ARDS diagnosis *among critical care physicians*, particularly in the interpretation of chest imaging^20^. Since chest images were unavailable, we relied on radiologist reports, using previously developed Berlin Definition-based inclusion and exclusion language to guide critical care physicians and reduce interrater disagreement^8^. This language also facilitated NLP processing for our ML approach, improving the signal-to-noise ratio in model development. While our approach mitigates variability from physician labeling of the reports, it assumes that radiologists—as the most trained in image interpretation—provide the most reliable clinical terminology description of an image. This is a common assumption, since radiologist reports are currently used to label images for computer vision models^39^. However, radiologists may also disagree in their interpretations^40,41^, which could limit our pipeline’s reliability since we had access to only one report per imaging study. To enhance reliability, future work could incorporate computer vision for direct chest image interpretation when available, using reports as a fallback. Integrating imaging will also require careful preprocessing to remove clinically irrelevant artifacts^5^, and a labeling process involving at least three radiologists, ideally assigning confidence ratings rather than binary classifications^42^.

Nevertheless, we examined the relationship between interrater disagreement and the Bilateral Infiltrates (BI) model’s confidence. Surprisingly, lower disagreement among our raters correlated with lower model confidence in a “yes” prediction, while higher disagreement occurred when the model was highly confident in “yes.”. Considering the results we show in Figure 3d, we speculate this may stem from differences in how physician raters labeled reports. Comparing individual raters to the BI model (Fig. S7), we found that the internal medicine physician rater had the highest rate of disagreement with the model when the model was highly confident in the “yes” choice. In contrast, the critical care physician rater had the highest rate of disagreement with the model when the model was most uncertain. This suggests the internal medicine physician was more cautious in labeling reports as positive, having adjudicated 196 reports as positive compared to 217 by the critical care physician.

Another potential limitation of our study is the absence of lung ultrasound data. This modality was included in the 2024 Berlin Definition update^37^, later than when data collection for the cohorts studied occurred. Nonetheless, our data includes chest X-ray and CT scan reports, which align with standard clinical practice at study institutions. Notably, the 2024 update does not equate lung ultrasound with these modalities but recommends it only when X-ray or CT is unavailable, such as in low-resource settings^37,43^. Thus, our pipeline may be less applicable in such settings.

Another limitation of our approach is the adjudication of ARDS risk factors. We faced significant challenge in implementing machine learning for attending physician notes due to the lack of clear inclusion/exclusion language. In addition, we had only 744 labeled notes for sepsis, the most common ARDS risk factor, compared to over 12,000 labeled chest imaging reports. Fortunately, a standard regex approach performed well in practice, with high accuracy and few false negatives.

Another limitation of our study is the relatively small number of medical centers and time periods from which we obtained data. While our approach proved robust across different models and datasets for adjudicating bilateral infiltrates, the same level of validation is lacking for risk factor determination and echocardiogram report adjudication. Implementing our pipeline in additional centers may require new regex patterns for echocardiogram reports, though the standardized nature of the required information mitigates this concern.

In summary, this study presents a computational pipeline for automating the Berlin Definition in retrospective adjudication of ARDS. Our pipeline performs excellently on both the development cohort and a subset of MIMIC-III encounters, highlighting the potential of computational techniques to address ARDS under-recognition. Future work will focus on clinical implementation.

## Supporting information

All supplementary information referenced in main text

## Acknowledgements

The authors thank Catherine Gao for insightful discussions and suggestions.

## Author Contributions

**FM -** Methodology, Software, Validation, Data Curation, Writing – Original Draft, Writing – Review & Editing, Visualization. **HAL -** Methodology, Software, Validation, Data Curation, Writing – Review & Editing. **HTN -** Software, Validation, Data Curation. **MB -** Methodology, Validation, Data Curation, Writing – Review & Editing. **FX -** Methodology, Software, Validation, Data Curation, Writing – Review & Editing, Visualization. **JK -** Data Curation. **ELC** - Data Curation, Writing – Review & Editing. **CHW -** Conceptualization, Methodology, Validation, Data Curation, Resources, Writing – Original Draft, Writing – Review & Editing, Visualization, Supervision, Project Administration, Funding Acquisition. **LANA -** Conceptualization, Methodology, Software, Validation, Formal Analysis, Resources, Writing – Original Draft, Writing – Review & Editing, Visualization, Supervision, Project Administration, Funding Acquisition.

## Data Availability

MIMIC-III is available on PhysioNet (https://doi.org/10.13026/C2XW26). Data from Hospital A (2013), Hospital A (2016), and Hospital B (2017-18) are IRB-protected. Upon publication, we will only release de-identified data from the Hospital (2013) cohort needed to reproduce Figure 8 at the ARCH repository hosted by Northwestern University (https://arch.library.northwestern.edu).

## Code Availability

The Python code to reproduce the reported results will be made available upon publication at the Amaral lab GitHub repository (https://github.com/amarallab/ARDS_diagnosis). These include the SQL scripts needed to pull the MIMIC (2001-12) cohort from MIMIC-III.

