## Supplementary material for "Open-source computational pipeline automatically flags instances of acute respiratory distress syndrome from electronic health records": All supplementary information referenced in main text

### A note on the calculation of $PaO_2/FiO_2$ (PF) ratios

For Hospital A (2013), we had previously collected all $FiO_2$, $PaO_2$, and PEEP values available for each encounter[8]. For some $PaO_2$ entries, there was a simultaneous $FiO_2$ and/or PEEP. However, for most encounters, we assumed that a $FiO_2$ or PEEP entry in the EHR remained the same until the next value entry; for example, if a $FiO_2$ of 40% was entered, we assumed that the $FiO_2$ the patient received was 40% until the next $FiO_2$ entry. As a result, the PF ratio was calculated based on the $FiO_2$ and PEEP that were the closest preceding entries for any $PaO_2$ entry.

The procedure for MIMIC (2001-12) was as follows: for each $PaO_2$ value, find its closest preceding value of $FiO_2$, and assign the timestamp of $PaO_2$ as the resulting PF ratio timestamp; in addition, for each $FiO_2$ value, find its closest preceding value of $PaO_2$, and assign the timestamp of $FiO_2$ as the PF ratio timestamp.

These calculations were performed prior to physicians performing retrospective reviews for both Hospital A (2013) and MIMIC (2001-12), and prior to data entering the adjudication pipeline. Therefore, both the human raters and the pipeline adjudicated each Berlin Definition component, and the overall ARDS label, using the same information.

### Finding documentation rate on MIMIC (2001-12)

Attending physician notes from the time MIMIC-III encompasses (2001-2012) may have used the “acute lung injury” nomenclature for what is now classified as mild ARDS. Our reviewers checked all MIMIC (2001-12) attending physician notes for “acute lung injury” or “ALI.” Only eight notes from two patients had these phrases. Interestingly, all these notes also contained “ARDS.” As a result, including the ALI term in the search did not change the ARDS documentation for MIMIC (2001-12).

**Figure S1. Most chest imaging reports from MIMIC (2001-12) occur within 15 hours of a PF ratio entry ≤ 300 mm Hg.** The graph shows a distribution of absolute time differences between the timestamps of chest imaging reports, and the timestamps of PF ratio ≤ 300 mm Hg entries. As such, reports could have been issued before or after the PF ratio entry.

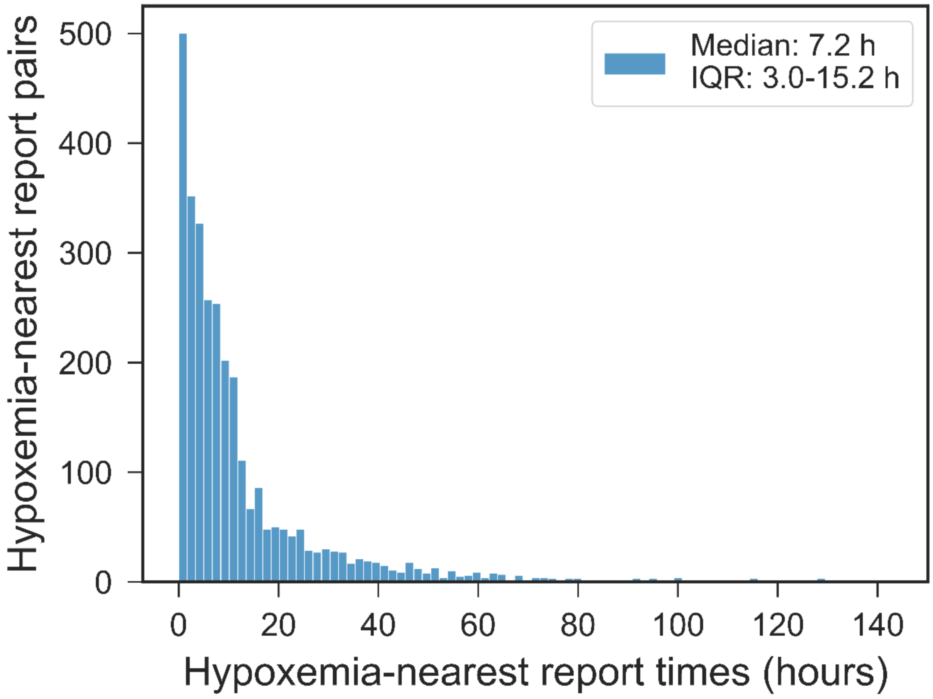

**Figure S2. SHAP value feature importances for Bilateral Infiltrates Model display concordance with inclusion and exclusion language.** Each word or set of words is sorted in descending order of mean absolute SHAP value, which measures each word's impact and contribution to XGBoost's output probabilities. Words like 'edema' and 'bilateral' are considered "inclusion language", therefore increased counts of these words move output probabilities by the Bilateral Infiltrates Model closer to 1. Conversely, 'atelectasis' is mostly considered "exclusion language", so increased counts of this word reassuringly result in the Bilateral Infiltrates Model decreasing the output probabilities toward 0.

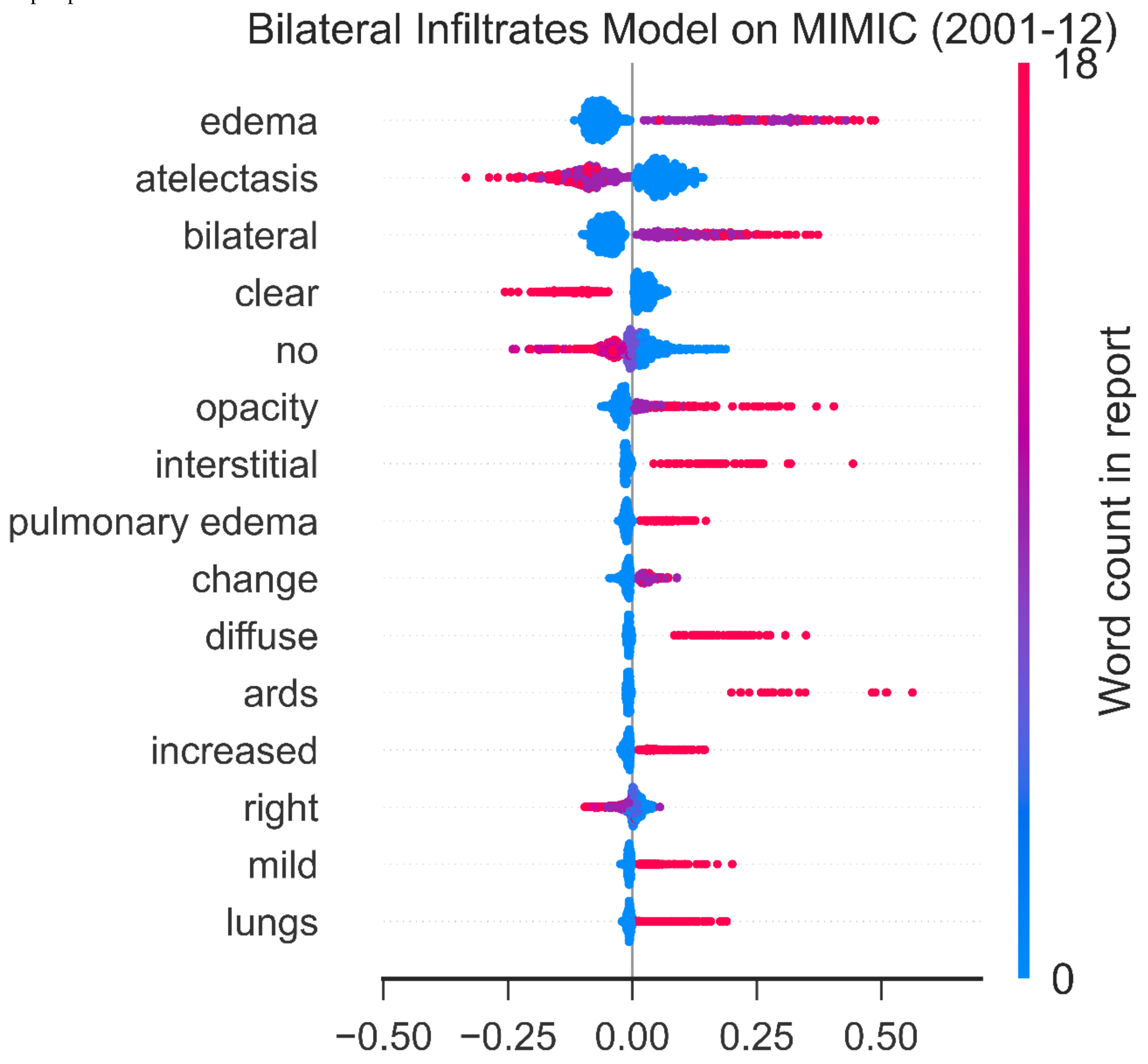

**Figure S3. Evaluation of XGBoost's robustness and generalization performance to Hospital B (2017-18) and MIMIC (2001-12) cohorts.** We show AUROCs with bootstrapped 95% confidence intervals as error bars for XGBoost models trained on **a)** chest imaging reports from Hospital A (2013), and **b)** chest imaging reports from Hospital A (2016).

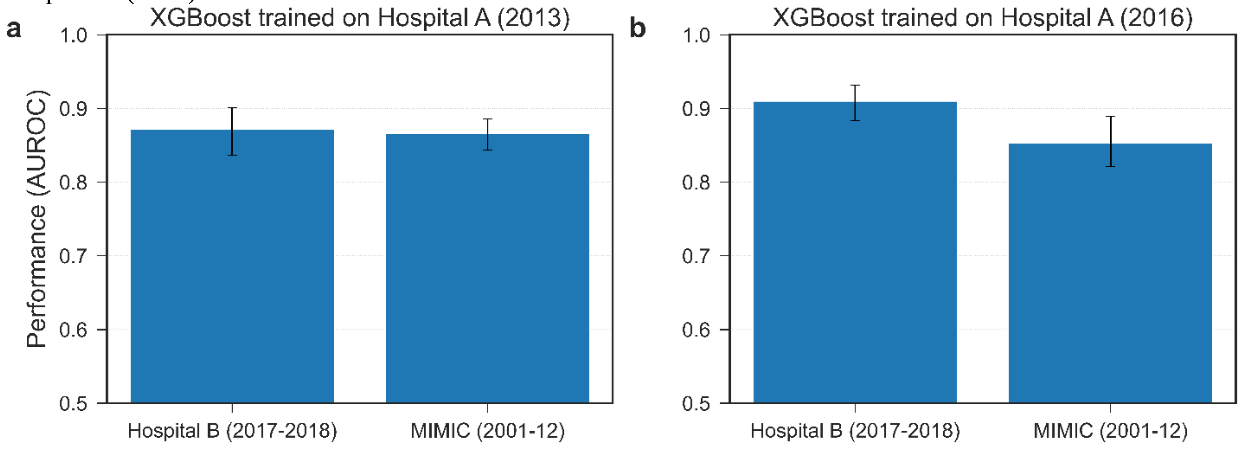


**Figure S4. Performance of the Bilateral Infiltrates Model at the encounter level.** ROC curve for the Bilateral Infiltrates Model tested on a single, randomly chosen chest imaging report per MIMIC (2001-12) encounter (100 encounters total). Error band shows 95% confidence interval for estimate of the mean ROC obtained using bootstrapping, which was obtained by repeating the random pick of chest imaging reports 100 times.

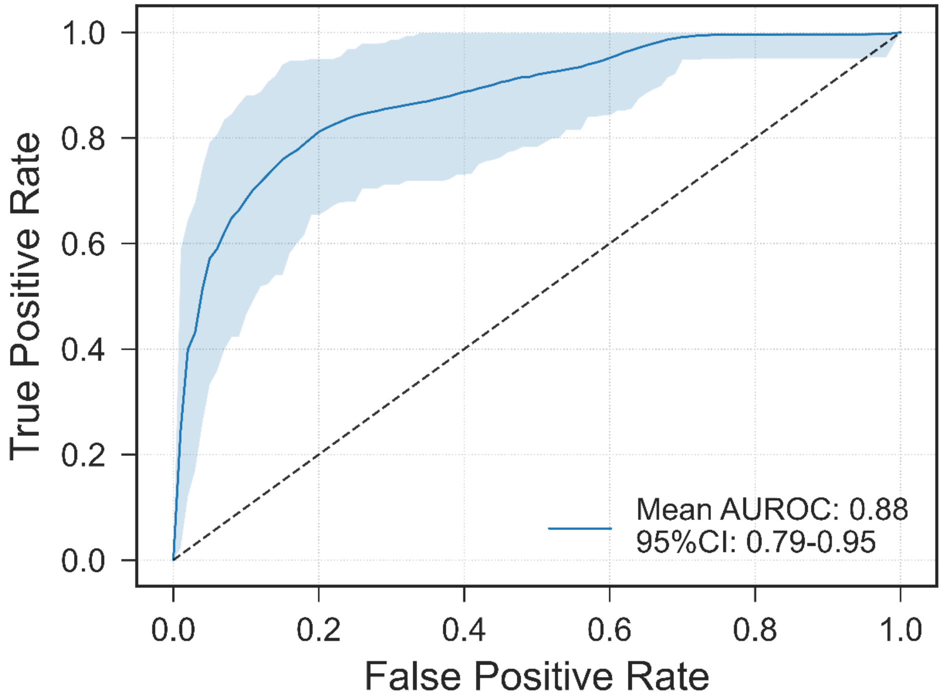

**Table S1. Exploration of alternative probability thresholds to binarize probabilities from the Bilateral Infiltrates (BI) Model when applied to chest imaging reports from MIMIC (2001-12).** We show probability thresholds (or ranges of probability thresholds) that optimize the accuracy, $F_1$ score, and Youden's J statistic, respectively. The optimal values are in boldface. Since optimal values for false positive rate, false negative rate, precision (or positive predictive value), and negative predictive value could be achieved with adjudicating no report, or all reports, with bilateral infiltrates, we simply boldface the best values for these metrics among the three thresholds considered.

| PROBABILITY THRESHOLD | FALSE NEGATIVE RATE | FALSE POSITIVE RATE | PRECISION | NEGATIVE PREDICTIVE VALUE | ACCURACY | $F_1$ SCORE | YOUDEN'S J STATISTIC |
|---|---|---|---|---|---|---|---|
| **84.7%** | 43.8% | **3.7%** | **81.3%** | 88.5% | **0.874** | 0.665 | 0.525 |
| **49.7%-49.8%** | 23.0% | 13.9% | 61.4% | 92.9% | 0.841 | **0.683** | 0.631 |
| **47.2%-47.9%** | **22.1%** | 14.5% | 60.6% | **93.1%** | 0.838 | 0.681 | **0.634** |

**Regular expression list 1: Risk factor/heart failure language capturing**

Regular expression approaches are specific to the text of interest. See the main text for which of these regular expression patterns were implemented in the pipeline. This is a list of all regular expressions developed during the study:

- **Pneumonia:** (?<!\w)(?:PCPpneumonia|pneumonia|Pneumonia|PNEUMONIA|pneumoniae|pneunonia|pneunoniae|pnuemonia|bronchopneumonia|parapneumonic|PNA|CAP|VAP|HAP|HCAP|hcap|infection|abx|PCP)(?!\w)
- **Aspiration:** (?i)(?<!\w)(?<!possibility\sof\s)(?<!\(\?)(?<!no\s{4}e\/o\s)(?<!unclear\sif\sthis\sis\s)(?<!cannot\srule\sout\s)(?<!risk\sfor\s)(?<!risk\sof\s)(?<!\?\s)(?<!cover\sfor\s)(?<!no\switnessed\s)(?:aspiration|aspirating)(?!\svs)(?!\svs.)(?!\?)(?!\ss\/p\sR\smainstem\sintubation)(?!\sprecautions)(?!\sand\sdrainage)(?!\w)
- **Inhalation:** (?i)(?<!\w)(?: inhaled\sswimming\spool\swater|inhalation\sinjury)(?!\w)
- **Pulmonary contusion:** (?i)(?<!\w)(?:pulmonary|pulmoanry)\s+(?:contusion|contusions)(?!\w)
- **Vasculitis:** (?i)(?<!\w)(?<!\?\s)(?<!less\slikely\s)(?:pulmonary\svasculitis|vasculitis)(?!\slabs)(?!\sworkup)(?!\sand\scarcinomatosis\sis\sless\slikely)(?!\shighly\sunlikely)(?!\sless\slikely)(?!\w)
- **Drowning:** (?i)(?<!\w)(?:drowned|drowning)(?!\w)
- **Sepsis:** (?i)(?<!\w)(?:sepsis|urosepsis|septiuc|septic|ssepsis|sseptic|spetic)(?!\w)
- **Shock:** (?i)(?<!\w)(?:shock|shocks|schock)(?!\w)
- **Overdose:** (?i)(?<!\w)(?:overdose|drug\soverdose)(?!\w)
- **Trauma:** (?i)(?<!\w)(?<!OGT\s)(?:trauma|traumatic|barotrauma|barotraumatic)(?!\w)
- **Pancreatitis:** (?i)(?<!\w)pancreatitis(?!\w)
- **Burns:** (?i)(?<!\w)(?:burn|burns)(?!\w)
- **TRALI:** (?i)(?<!\w)(?:TRALI|(?i)transfusion(?:-|\s)related\sacute\slung\sinjury|(?i)transfusion(?:-|\s)associated\sacute\slung\s injury)(?!\w)
- **ARDS:** (?i)(?<!\w)(?:ards|acute\srespiratory\sdistress\ssyndrome|acute\slung injury|ali|ardsnet|acute\shypoxemic\srespiratory\sfailure)(?!\w)
- **Pregnant:** (?i)(?<!\w)(?:IUP|G\dP\d)(?!\w)
- **CHF:** (?i)(?<!\w)(?<!h\/o\s)(?:congestive\sheart\sfailure|chf|diastolic\sHF|systolic\sHF|heart\sfailure|diastolic\sdysfunction|LV\sdysfunction|low\scardiac\soutput\ssyndrome|low\scardiac\soutput\ssyndrom|low\scardiac\souput\ssyndrome|low\sCO\sstate)(?!\swith\spreserved\sef)(?!\swas\sanother\spossible\sexplan)(?!\w)
- **Cardiogenic:** (?i)(?<!\w)(?<!no\se\/o\sobstructive\sor\s)(?<!versus\s)(?<!rule\sout\s)(?<!ruled\sout\s)(?<!less\slikley\s)(?<!w\/o\sevidence\ssuggestive\sof\s\s)(?<!non\s)(?<!less\slikely\s)(?<!not\slikely\s)(?<!unlikely\sto\sbe\s)(?<!no\sclear\sevidence\sof\sacute\s)(?<!non-)(?<!than\s)(?<!no\sevidence\sof\s)(?:cardiogenic|cardigenic|cardiogemic|cardiac\spulmonary\sedema|cardiac\sand\sseptic\sshock|Shock.{1,15}suspect.{1,15}RV\sfailure)(?!\s\(not\slikely\sgiven\sECHO\sresults\))(?!\sshock\sunlikely)(?!\svs\.\sseptic)(?!\scomponent\salthough\sSvO2\snormal)(?!\w)
- **Non-cardiogenic:** (?i)(?<!\w)(?:non(?:-|\s)cardiogenic|noncardiogenic|non(?:-|\s)cardigenic|noncardigenic)(?!\w)

- **Palliative:** (?i)(?<!\w)(?:palliative\scare|comfort\scare|withdraw\scare|comfort\salone|withdraw\ssupport\sin\sfavor\sof\spalliation)(?!\w)
- **Cardiac arrest:** (?i)(?<!\w)(?:arrest|cardiorespiratory\sarrest)(?!\w)
- **Dementia:** (?i)(?<!\w)dementia(?!\w)
- **Stroke:** (?i)(?<!\w)(?:stroke|strokes|cerebellar hemorrhage|intracerebral hemorrhage|BG hemorrhage|cva|cerebrovascular accident|cefrebellar infarcts\/basilar stenosis)(?!\w)
- **Alcohol:** (?i)(?<!\w)(?:alcohol\swithdrawal|dts|dt"s|dt|alcohol\sdependence|alcohol\sabuse|etoh\sabuse|etoh\swithdrawal|etoh\swithdrawl|etoh\sw\/drawal|delirium\stremens)(?!\w)
- **Seizure:** (?i)(?<!\w)(?<!no e/o subclinical\s)(?:seizure|seizures)(?!\w)
- **AMI:** (?i)(?<!\w)(?:ami|acute\s myocardial\s ischemia|acute\s myocardial\s infarction|myocardial\sinfarction|nstemi|non-st\s elevation\s mi|stemi|st\s elevation\s mi|acute\smi)(?!\w)

**Figure S5. Capturing performance of regular expressions listed in Regular expression list 1.** Most regular expressions developed capture 100% of the attending physician notes from Hospital A (2013) labeled as “yes” for each risk factor. ‘Sepsis’ and ‘shock’ are the most prevalent risk factors for ARDS after pneumonia. ‘Cardiogenic’ and ‘congestive heart failure’ are the most prevalent heart failure keywords in attending physician notes from Hospital A (2013).

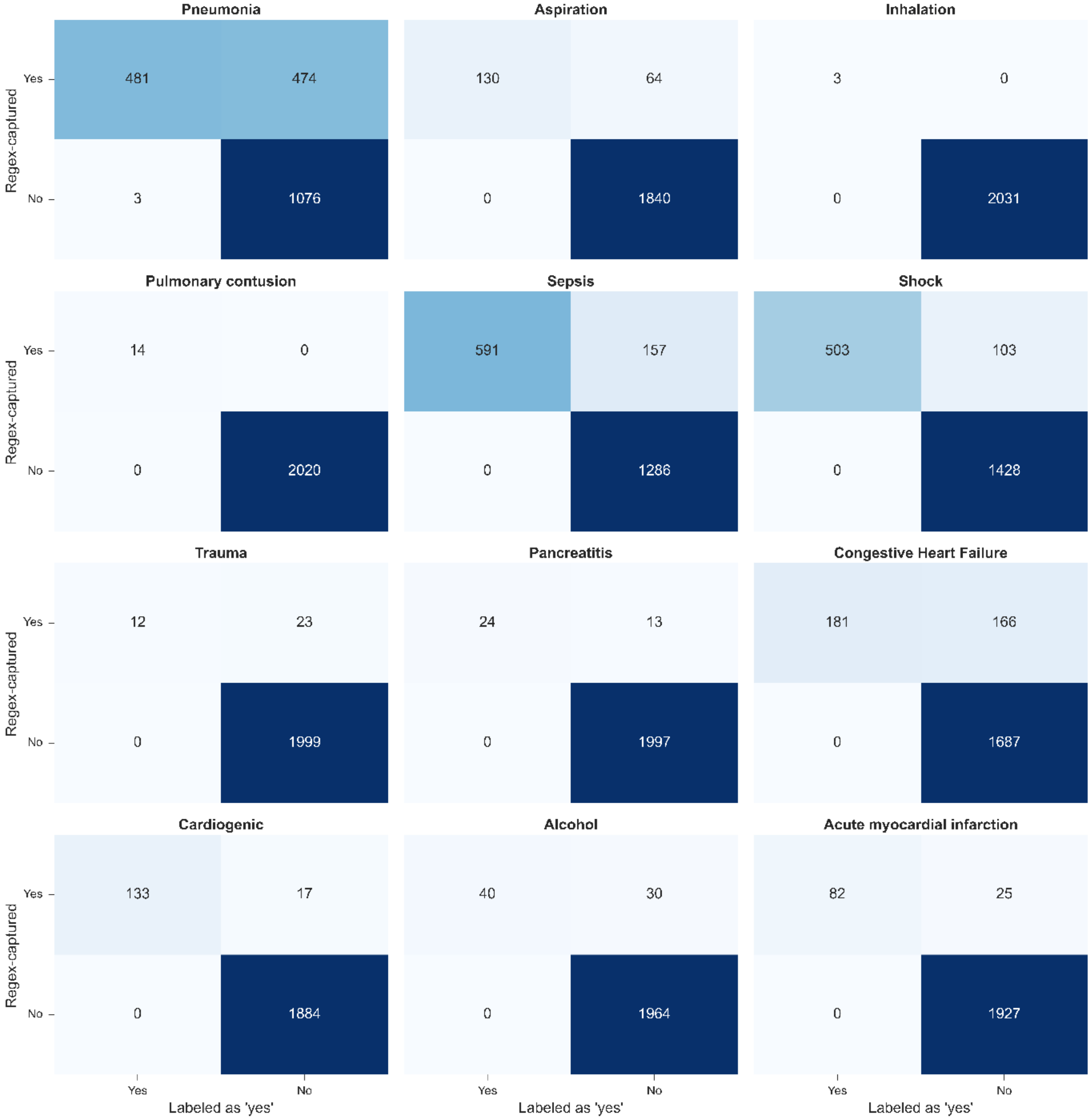

**Regular expression list 2: "Exclusion" key words or phrases**
We used the following key words or phrases to adjudicate a note as **negative** for a risk factor or heart failure criterion regardless of their being captured by regular expressions from list 1:

- **Sepsis:** 'r/o sepsis', 'no longer in', 'sepsis or cardiogenic shock', 'cardiogenic vs septic', 'potential for septic shock', 'cardiac vs septic', 'searching for evidence of', 'shock mixed cardiogenic/vasodilatory', 'sepsis vs cardiogemnic', 'shock-septic vs cargdiogenic', 'shock-septic vs cardiogenic', 'severe sepsis resolved', 'now off low dose vasopressor', 'shock-septic vs hypovolemic resolved', 'septic schock-resolved', 'shock-septic vs hypovolemic', 'septic shock off pressors', 'cannot rule out septic/vasodilatory shock', 'previously vasoactive support for sepsis', 'billiary sepsis', 'septic shock secondary to esbl bacteremia', 'c/b septic joints', 'septic shock due to pseudomonas bacteremia', 'no evidence of hemorrhage or sepsis', 'no evidence of ongoing hemorrhage or sepsis', 'admitted with septic shock about months ago', 'history of aspergillus pneumonia/sepsis', 'mssa bactermia septic shock resolved', 'hypotension/sepsis vs hypercoaguable', 'septic shock ards copd exacerbation hcap bacteremia', 'septic shock found to have klebsiella bacteremia', 'suspected sepsis', 'septic emboli syndrome', 'takotsubo possible', 'takatsubo with possible', 'septic shock suspect recurrent takatsubo's', 'without active hemorrhage or sepsis', 'w/u for sepsis underway', 'also concern for sepsis', 'cytopenias likely due to sepsis', 'sedation sepsis', 'septic shock with picture', 'no signs of sepsis at this time', 'potentially sepsis', 'does not have septic shock', 'sepsis unlikely', 'no evidence of sepsis', 'does not have sig signs/sxs infection or sepsis', 'no source of sepsis', 'h/o urosepsis'.
- **Shock:** 'is no longer in septic shock', 'chest compressions or shocks', 'shock now resolved', 'potential for septic', 'septic shock off pressors', 'septic shock with picture', 'no longer in shock', 'septic shock-resolved', 'septic shock due to pseudomonas bacteremia', 'shock has resolved', 'septic shock source uncertain', 'not in shock', 'weaned off pressors', 'septic shock due to e-coli bactermia resolved', 'most likely distributive liver failure vs sepsis', 'shock--improving', 'terminated by shock', 'icd interrogation reported two shocks', 'unlikely to be cardiogenic shock', 'cpr/shocks', 'shocks before rosc', 'underwent cardiopulmonary resuscitation and', 'now off low dose vasopressor requirement', 'schock-resolved', 'vs hypovolemic resolved', 'cannot rule out septic/vasodilatory', 'obstructive shock due to pe and septic', 'no operative intervention recommended except in shock situation'.

**Figure S6. Performance of exclusion keywords and key phrases in reducing false positive rates.** Language to exclude notes successfully reduced false positives in sepsis from 157 to 48, and in shock from 103 to 55 when applied to attending physician notes from Hospital A (2013). We did not include other risk factors in this approach due to our inability to find exclusion words or phrases that would decrease false positives while preserving the true positives.

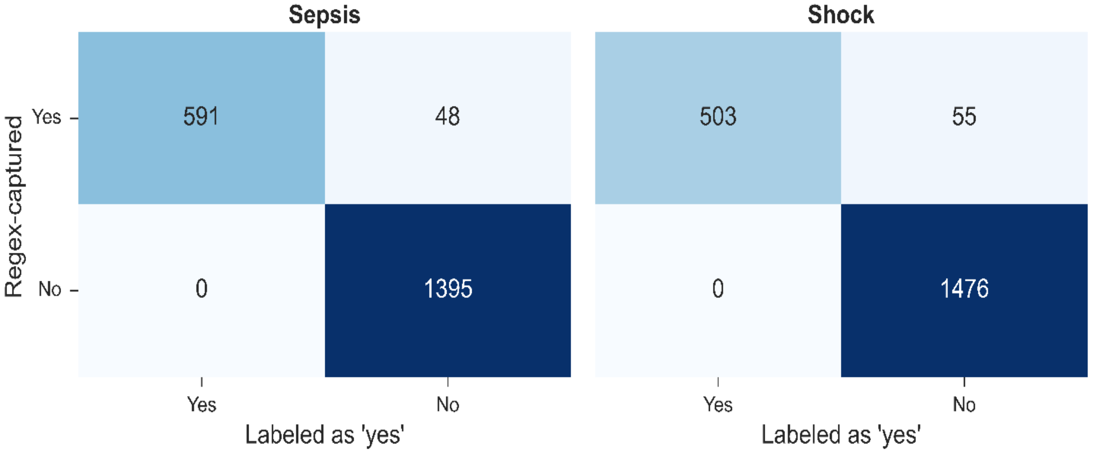

**Table S2.** Number of attending physician notes that were reviewed for each risk factor/heart failure keyword, labeled as yes vs. no, and also regex-matched.

| Risk factor or heart failure | Number of notes labeled | % labeled yes | Number of notes regex-matched | % yes among regex-matched |
|---|---|---|---|---|
| Sepsis | 744 | 79% | 748 | 79% |
| Pneumonia | 636 | 76% | 955 | 51% |
| Shock | 604 | 83% | 606 | 83% |
| Congestive Heart Failure | 254 | 71% | 347 | 52% |
| Aspiration | 221 | 59% | 194 | 67% |
| ARDS | 190 | 73% | 266 | 52% |
| Cardiogenic | 176 | 76% | 150 | 89% |
| Stroke | 156 | 44% | 156 | 44% |
| Acute myocardial infarction | 143 | 57% | 107 | 77% |
| Seizure | 129 | 33% | 129 | 33% |
| Cardiac arrest | 105 | 98% | 139 | 74% |
| Alcohol | 81 | 49% | 70 | 57% |
| Inhalation | 70 | 4% | 3 | 100% |
| Trauma | 49 | 24% | 55 | 22% |
| Pancreatitis | 36 | 67% | 37 | 65% |
| Palliative | 21 | 76% | 24 | 67% |
| Pulmonary contusion | 14 | 100% | 14 | 100% |
| Dementia | 13 | 100% | 16 | 81% |
| Pregnant | 11 | 100% | 12 | 92% |
| Non-cardiogenic | 7 | 43% | 7 | 43% |
| Burn | 7 | 0% | 4 | 0% |
| TRALI | 6 | 67% | 6 | 67% |
| Vasculitis | 4 | 100% | 4 | 100% |
| Drowning | 3 | 100% | 3 | 100% |
| Overdose | 2 | 100% | 2 | 100% |

**Table S3. Exploration of alternative probability thresholds to binarize probabilities from the Pneumonia Model on MIMIC (2001-12) attending physician notes.** We show probability thresholds (or ranges of probability thresholds) that optimize the accuracy, $F_1$ score, and Youden's J statistic, respectively. The optimal values are in boldface. Since optimal values for false positive rate, false negative rate, precision (or positive predictive value), and negative predictive value could be obtained by adjudicating pneumonia for all, or no notes, we simply boldface the best values for these metrics among the thresholds considered.

| Probability threshold | False negative rate | False positive rate | Precision | Negative predictive value | Accuracy | $F_1$ score | Youden's J statistic |
|---|---|---|---|---|---|---|---|
| 22.7%-22.8% | 27.8% | 5.8% | 86.7% | 86.7% | **0.867** | 0.788 | 0.665 |
| 24.2%-24.7% | 28.1% | **5.6%** | **87.0%** | 86.6% | **0.867** | 0.787 | 0.663 |
| 19.3%-19.8% | 24.8% | 7.7% | 83.5% | 87.8% | 0.865 | **0.791** | 0.675 |
| 20.0%-20.6% | 25.2% | 7.5% | 83.8% | 87.6% | 0.865 | **0.791** | 0.673 |
| 18.6% | **23.7%** | 8.8% | 81.7% | **88.1%** | 0.861 | 0.789 | **0.675** |
| 19.3%-19.8% | 24.8% | 7.7% | 83.5% | 87.8% | 0.865 | **0.791** | **0.675** |

**Regular expression list 3: Echocardiogram reports**

Regular expression approaches are specific to the text of interest. For flagging relevant metrics in echocardiogram reports, we used a two-pronged approach: prefix regular expressions, and suffix regular expressions:

- **Left ventricular ejection fraction:**
    - ***Prefix***: (?i)(?:lv\s+ejection\s+fraction|left\s+ventricular\s+ejection\s+fraction|lvef|left\s+ventricular\s+ef|lvef\s+is|left\s+ventricle\s+ejection\s+fraction\s+is|lv\s+ejection\s+fraction\s+is)
    - ***Suffix***: \D{0,20}(\d{1,3}|\d{1,2}\s*-\s*\d{1,3})-{0,1}\s*%
- **Cardiopulmonary bypass:**
    - ***Prefix***: (?i)cardiopulmonary\s+bypass
    - ***Suffix***: (?!\s*N\/A|\s*Patient\s+was\s+not\s+placed\s+on\s+cardiopulmonary\s+bypass|\s*NA)
- **Left atrial dimension:**
    - ***Prefix***: (?i)(?:la\s+diameter|la\s+dimension)
    - ***Suffix***: \D{0,25}(\d\.\s*\d)\s*(?:cm|centimeter)
- **Left atrial volume index:**
    - ***Prefix:*** (?i)(?:la\s+volume| LA\s+Vol\s+BP\s+A/L\s+Index)
    - ***Suffix:*** *.?(?:ml)?*.?(\d+.\s*\d+)\s+(?:(?=ml/m)|(?=ml\s+per\s+square\s+meter))
- **Left ventricular hypertrophy:**
    - ***Prefix***: (?i)(?:(?<!borderline )(?:left\s+ventricular|lv|lv\s+concentric)\s*hypertrophy|(?<!borderline )LVH)
    - ***Suffix***: No suffix
- **Diastolic dysfunction:**
    - ***Prefix***: (?i)(?:grade\s*ii|grade\s*iii)
    - ***Suffix***: .{0,30}\s*?(?=diastolic\s+dysfunction)

**Table S4. Assessment of regular expression accuracy in capturing values from echo reports.** In contrast to chest imaging reports or attending physician notes, we lacked access to echo reports labeled or labeled for the specific value of the factors. To validate our regex-matching procedure for each objective heart failure criterion (which included numeric or text values), we took a subset of 10% of echo reports for manual annotation/validation. Specifically, to validate left ventricular ejection fraction, left atrial diameter and left atrial volume index, we took 10% of echo reports that were flagged for that factor and had a corresponding value, and another 10% sample of echo reports flagged for that factor but did not have a value. In addition, we took a 10% sample among all echo reports to validate cardiopulmonary bypass, left ventricular hypertrophy, and diastolic dysfunction. Next, we computed the Hamming distance between the regex-found values and the manually validated values as a metric of concordance between both value arrays. In short, the Hamming distance is the fraction of corresponding entries that are different between both arrays. After a few regex-pattern development iterations, we find perfect agreement between our regex-matching approach and the validated values.

| **Objective Heart Failure factor** | Value/ No Value | # of echo reports | Hamming distance | 1-Hamming distance |
|---|---|---|---|---|
| Left ventricular ejection fraction | Value | 93 | 0 | 1 |
| | No Value | 8 | 0 | 1 |
| Cardiopulmonary bypass | na | 101 | 0 | 1 |
| Left atrial diameter | Value | 50 | 0 | 1 |
| | No Value | 48 | 0 | 1 |
| Left atrial volume index | Value | 40 | 0 | 1 |
| | No Value | 60 | 0 | 1 |
| Left ventricular hypertrophy | na | 101 | 0 | 1 |
| Grade II or III diastolic dysfunction | na | 101 | 0 | 1 |

### Adjudication of ARDS on Hospital A (2013): Objective heart failure assessment

Of the 102 encounters entering heart failure assessment, 14 were excluded due to having BNP >100 pg/mL. Of the remaining 88 encounters, 9 were discarded due to showing LVEF < 40% in their echocardiogram reports. Then, 34 encounters had evidence of cardiopulmonary bypass, while 45 encounters did not show evidence of cardiopulmonary bypass. Finally, out of the 45 encounters remaining, 6 had two of left atrial enlargement, left ventricular hypertrophy and/or diastolic dysfunction. This resulted in 44 encounters who did not show evidence of heart failure through the objective assessment and were adjudicated as ARDS by the pipeline.

### Adjudication of ARDS on MIMIC (2001-12): Objective heart failure assessment

Of the 13 encounters entering heart failure assessment, 0 were excluded due to having BNP >100 pg/mL. Of the remaining 13 encounters, 1 was discarded due to showing LVEF < 40% in their echocardiogram reports. Then, 0 encounters had evidence of cardiopulmonary bypass. Finally, out of the 12 encounters remaining, 1 had two of left atrial enlargement, left ventricular hypertrophy and/or diastolic dysfunction. This resulted in 11 encounters who did not show evidence of heart failure through the objective assessment and were adjudicated as ARDS by the pipeline.

**Table S5. Impact of alternative probability thresholds to binarize Bilateral Infiltrates Model's probabilities on the ARDS computational pipeline's metrics for Hospital A (2013).** We show probability thresholds (or ranges of probability thresholds) that optimize the accuracy, $F_1$ score, and Youden's J statistic for the Bilateral Infiltrates Model individually (top three rows), and the entire ARDS adjudication pipeline (bottom row). The optimal values are in boldface. Since optimal values for false positive rate, false negative rate, precision (or positive predictive value), and negative predictive value could be done by adjudicating ARDS for all, or no encounters, we simply boldface the best values for these metrics among the thresholds considered. Note that the thresholds for the Pneumonia Model could not be explored since not every attending physician note from Hospital A (2013) was labeled for pneumonia.

| BI model threshold | Pneumonia Model threshold | FNR | FPR | Precision | NPV | Accuracy | $F_1$ | Youden's J |
|---|---|---|---|---|---|---|---|---|
| **48.6%-48.7%** | 50% | 4.3% | 13.6% | 77.0% | 97.7% | 0.894 | 0.854 | 0.821 |
| **27.1%-29.2%** | 50% | **2.9%** | 16.0% | 74.4% | **98.3%** | 0.882 | 0.842 | 0.811 |
| **49.9%-51.5%** | 50% | 4.3% | 13.3% | 77.5% | 97.7% | 0.896 | 0.856 | 0.824 |
| **57.4%-58.4%** | 50% | 4.3% | **12.4%** | **78.7%** | 97.7% | **0.902** | **0.864** | **0.834** |

**Table S6. Impact of alternative probability thresholds to binarize models' probabilities on the ARDS computational pipeline metrics's for MIMIC (2001-12).** We show probability thresholds (or ranges of probability thresholds) that optimize the accuracy, $F_1$ score, and Youden's J statistic for the Bilateral Infiltrates Model and Pneumonia Model individually (top three rows), and the entire ARDS adjudication pipeline (bottom row). The optimal values are in boldface. Since optimal values for false positive rate, false negative rate, precision (or positive predictive value), and negative predictive value could be done by adjudicating ARDS for all, or no encounters, we simply boldface the best values for these metrics among the thresholds considered.

| BI model threshold | Pneumonia Model threshold | FNR | FPR | Precision | NPV | Accuracy | $F_1$ | Youden's J |
|---|---|---|---|---|---|---|---|---|
| **84.7%** | 22.7%-24.7% | 12.9% | **10.1%** | **79.4%** | 93.9% | **0.89** | **0.831** | **0.770** |
| **49.7%-49.8%** | 19.3%-20.6% | **6.5%** | 17.4% | 70.7% | **96.6%** | 0.86 | 0.806 | 0.762 |
| **47.2%-47.9%** | 19.3%-19.8% | **6.5%** | 17.4% | 70.7% | **96.6%** | 0.86 | 0.806 | 0.762 |
| **84.7%-86.6%** | 16.0%-80.7% | 12.9% | **10.1%** | **79.4%** | 93.9% | **0.89** | **0.831** | **0.770** |

### Timing of ARDS adjudication for MIMIC (2001-12)

We conducted a sensitivity analysis limited to early chest imaging. We find that 37 out of the 41 encounters (90.2%) recognized by the automatic adjudication pipeline had a qualified hypoxemia event (PF ratio≤300 mm Hg and bilateral infiltrates recognized by the pipeline) within 48h of intubation. We then observe 39 (95.1%) encounters having at least one qualified hypoxemia episode within 72h of intubation. When considering the earliest episode of qualified hypoxemia post-intubation, we find the median time from intubation to earliest episode was 3.9 hours (IQR: 0.7h-21.8h), with two outlier encounters having the first episode after ~125 hours (~5 days) and ~355 hours (14 days) post-intubation.

**Figure S7. Rate of disagreement between physician raters and the Bilateral Infiltrates Model when adjudicating bilateral infiltrates on MIMIC (2001-12) chest imaging reports.** Error bars represent 95% confidence intervals. High confidence No corresponds with BI Model probabilities between 0% and 10%, Low confidence corresponds to probabilities between 10% and 90%, and High confidence Yes corresponds to probabilities between 90% and 100%. Within and Outside 48 h refers to whether the reports were within 48 h of a hypoxemic entry (PF ratio <=300 mm Hg) or not.

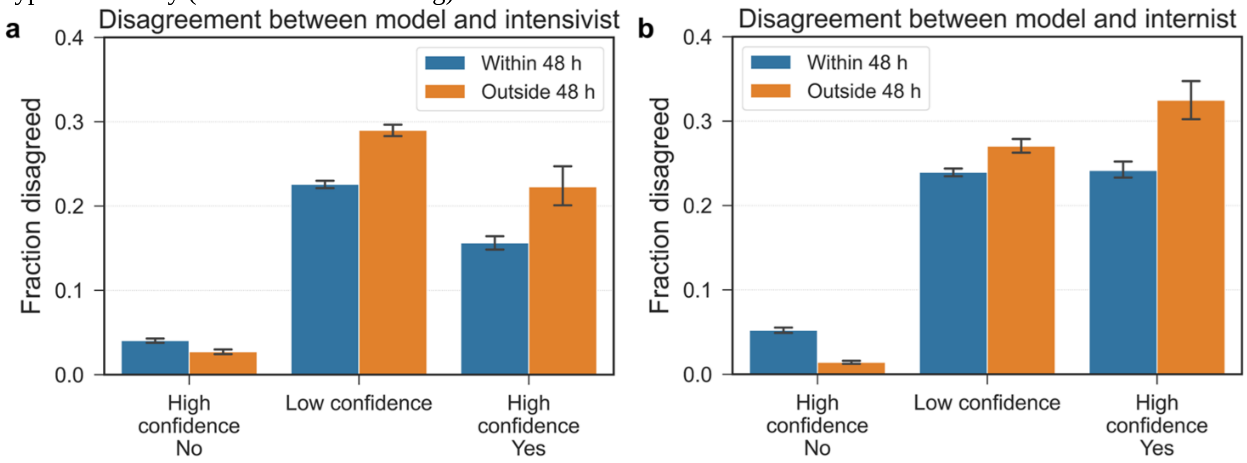
